# Gendered behaviors are associated with health beyond binary sex

**DOI:** 10.64898/2026.09.28.26364210

**Authors:** Olivier Parent, Maria McGuinness, Sophia Osborne, Matthew Danyluik, Alice Mukora, Daniela Quesada, Gabriel A. Devenyi, Armin Raznahan, Robert-Paul Juster, Mahsa Dadar, M. Mallar Chakravarty

## Abstract

Sex and gender are distinct but intertwined determinants of health. Both adherence to and deviation from gender norms carry consequences for health, yet gender’s contribution to disease risk beyond binary sex remains largely unexplored. As gender is organized around sex from its developmental origins, masculine and feminine health behaviors can be defined by how males and females behave collectively. Individuals vary in how closely they align with these norms, defining a biopsychosocial gender spectrum. Here, we used machine learning to reconstruct population-specific masculine and feminine behavioral norms, yielding a continuous Index of Gendered Life-domains (IGL) in 301,113 UK Biobank participants. In sex-stratified analyses of body and brain health phenotypes, masculine behavioral profiles were associated with greater physiological load and cardiometabolic disease risk, and feminine profiles with higher trait anxiety and musculoskeletal vulnerability. Sex–IGL incongruence predicted state-like psychiatric symptoms and severe mental health diagnoses. An intersectional approach revealed differences in gendered behaviors and their health associations across ethnic groups, demonstrating that gender measurement must be sensitive to sociocultural context to characterize health risk in diverse populations. These findings establish gender as a socioculturally embedded determinant of health whose health associations go beyond binary sex.

## Introduction

Sex and gender are often treated interchangeably in biomedical research, despite representing distinct constructs with different implications for health and well-being.^1^ Chromosomal sex is often defined as a binary variable, with exceptions such as sex chromosome aneuploidies. Gender extends beyond gender identity and encompasses gendered expectations, interindividual relations, and systemic interactions with institutional structures, all of which are shaped by local cultural contexts.^2^ While the study of sex as a modifier of health outcomes has gained important traction since the NIH Revitalization Act of 1993 mandated the inclusion of females in clinical research,^3^ the scarcity of gender measures in biomedical datasets has limited the examination of gender as a determinant of health.^1^ Because both adherence to and deviation from gender norms carry health implications,^4,5^ characterizing gender’s population-level health associations is essential for understanding epidemiological trends.

Measuring sociocultural gender as distinct from biological sex has proved challenging, since this is akin to isolating nurture from nature while complex behaviors likely emerge from their constant interactions.^6^ The sex/gender entanglement framework instead argues that both are intrinsically linked.^7^ In development, sex assigned at birth acts as a category around which gendered behaviors are organized, from social conduct to lifestyle and health habits. Children model sex-typed behavior from others who share their perceived sex category,^8^ then organize it into an internal schema.^9^ These internalized representations are foundational and continue to shape everyday behavior into adulthood.^10^ Gender norms and individual behavior are thus mutually constitutive, as masculine and feminine norms emerge from how males and females behave in aggregate across a population, which in turn guides how each individual behaves.^11^

From this theoretical and empirical background emerges the following conceptualization: masculine and feminine behaviors are those enacted more often by males and females, respectively, in a specific community. Each individual enacts different variations of masculine and feminine behaviors, and this variation collectively forms a continuous gender spectrum that complements binary sex. These dynamics apply to behavioral risk factors (e.g., substance use, diet, physical activity), which can differ in their gendered directions and importance across communities,^4^ thus providing a pathway through which socioculturally embedded gender operates as a determinant of health. Accordingly, throughout the manuscript, we use *sex* to designate binary chromosomal sex and *gender* to designate adherence to masculine and feminine behavioral norms shaped by biopsychosocial factors, distinct from gender identity or purely sociocultural influences on behavior.

We propose that these gender dynamics can be formally operationalized using data-driven gender scores in which binary sex is algorithmically classified from behavior.^12,13^ We introduce the Index of Gendered Life-domains (IGL), a data-driven continuous measure of gender derived using a sex classification machine learning model trained on behavioral and socioeconomic predictors in a large, population-scale dataset (UK Biobank; *n*=301,113). In our approach, population-level data inform the algorithm about which behaviors are normatively masculine or feminine. It then makes continuous predictions about how closely each individual’s behaviors align with these population-typical gendered patterns, yielding a gender measure positioned to be investigated as a determinant of health. Crucially, this approach does not *a priori* assign gendered directions or importance to behaviors, unlike questionnaire-based measures.^14^ This provides the necessary flexibility to capture the ever-changing nature of gendered norms across cultures and epochs under a unified framework. This is an essential feature, as the contents of masculine and feminine traits continuously evolve through time^15^ and vary across ethnicities^16^ and socioeconomic strata.^17^

Deploying the IGL in a population-scale cohort, we demonstrate that masculine and feminine health behavior norms are associated with distinct risk profiles for mental and physical health beyond binary sex. Critically, IGL-health associations vary substantially across ethnic communities, underscoring that gendered health risk cannot be understood outside specific sociocultural contexts. These findings establish gender as a measurable and consequential determinant of health beyond binary sex, fundamentally embedded in the sociocultural context in which gendered behaviors are expressed.

## Results

### A data-driven index of gendered life-domains

Using population-scale data from the UK Biobank (UKBB), comprising middle-aged and older adults (*n*=301,113; Table 1), we selected 79 variables representing diet, lifestyle, social, and socioeconomic factors to construct the Index of Gendered Life-domains (IGL; Methods; Supp. Table 1; Supp. Methods 1). We focused on behavioral and socioeconomic factors that are not fixed by biological sex. We trained CatBoost machine learning models to predict the genotyping-based chromosomal sex of participants from this variable set, capturing complex non-linear trends representing population-level gender norms (Fig. 1A; Supp. Figs. 1–2). How each individual aligned with these patterns was captured by the classification probability, the IGL, with values near 1 indicating a masculine IGL and values near 0 indicating a feminine IGL. This operationalization provides the basis to test gender as a determinant of health beyond chromosomal sex.

**Figure 1.**
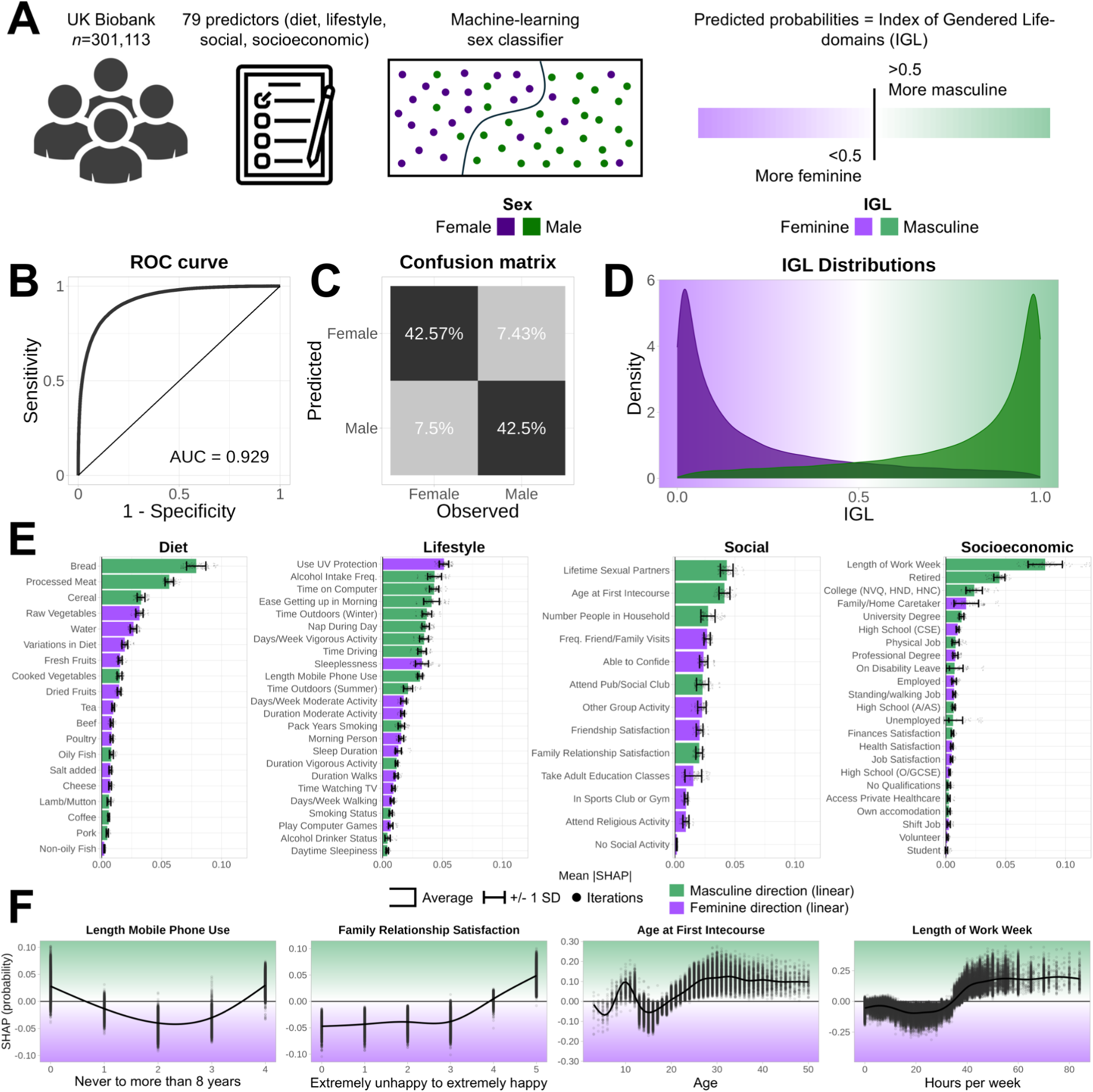
Index of gendered life-domains. **A)** CatBoost models were trained using five-fold cross-validation repeated 25 times to predict chromosomal sex from an array of predictors related to diet, lifestyle, social, and socioeconomic factors. The receiver operating characteristic (ROC) curve, area under the curve (AUC) values **(B)**, and confusion matrix **(C)** were calculated on averaged out-of-fold predictions, representing the Index of Gendered Life-domains (IGL). **D)** IGL distributions for females (dark purple) and males (dark green). Values below 0.5 indicate a more feminine IGL (light purple), while values above 0.5 indicate a more masculine IGL (light green). **E)** Mean absolute SHAP values were calculated for all 125 models and averaged, indicating variable importance. Error bars indicate +/- one standard deviation (SD). For visualization purposes, a linear direction was calculated with a Pearson correlation on the averaged SHAP plot and is indicated by a light purple (feminine) or light green (masculine) color. **F)** Examples of SHAP plots for variables that showed notable non-linear trends. For continuous variables, large positive outliers above the 99.9th percentile were excluded for visualization purposes only. Generalized additive models were fit to the data using a varying number of basis dimensions (equivalent to the number of unique values up to 15).

**Figure 2.**
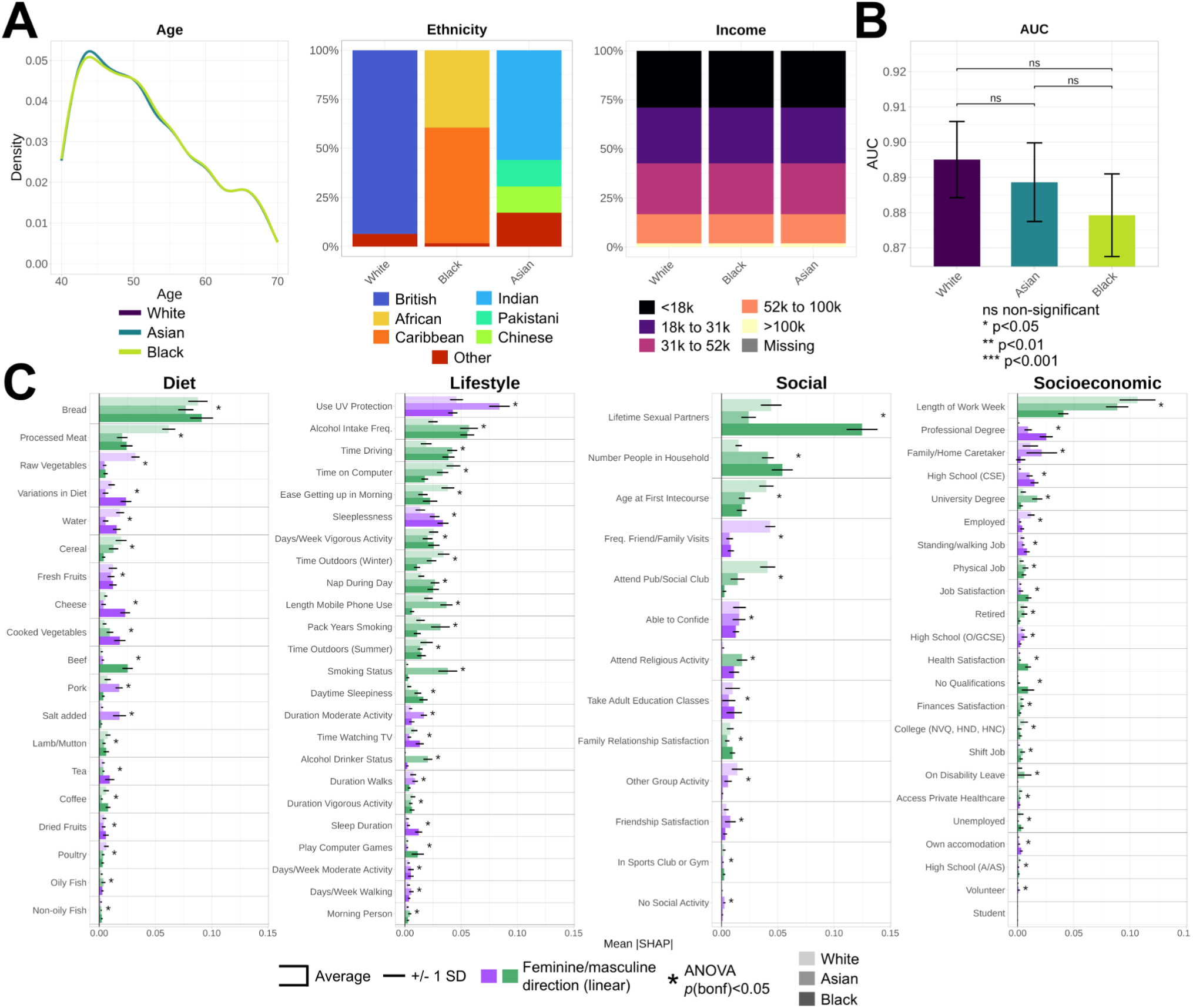
Gendered dynamics across ethnicities. IGL models were computed in ethnicity groups matched on age and household income. **A)** Distributions of age, ethnicity subgroup, and household income across ethnicities. **B)** AUC values across ethnicities with 95% confidence intervals, compared with unpaired DeLong tests. **C)** Variable importances across ethnicities. Error bars indicate +/- one standard deviation (SD). A linear direction was calculated with a Pearson correlation on the averaged SHAP plot and is indicated by a light purple (feminine) or light green (masculine) color. Statistical differences between groups for each variable were calculated using ANOVAs, *p*-values were adjusted with a Bonferroni correction, and significant effects at *p*<0.05 are indicated with an asterisk.

**Table 1.**
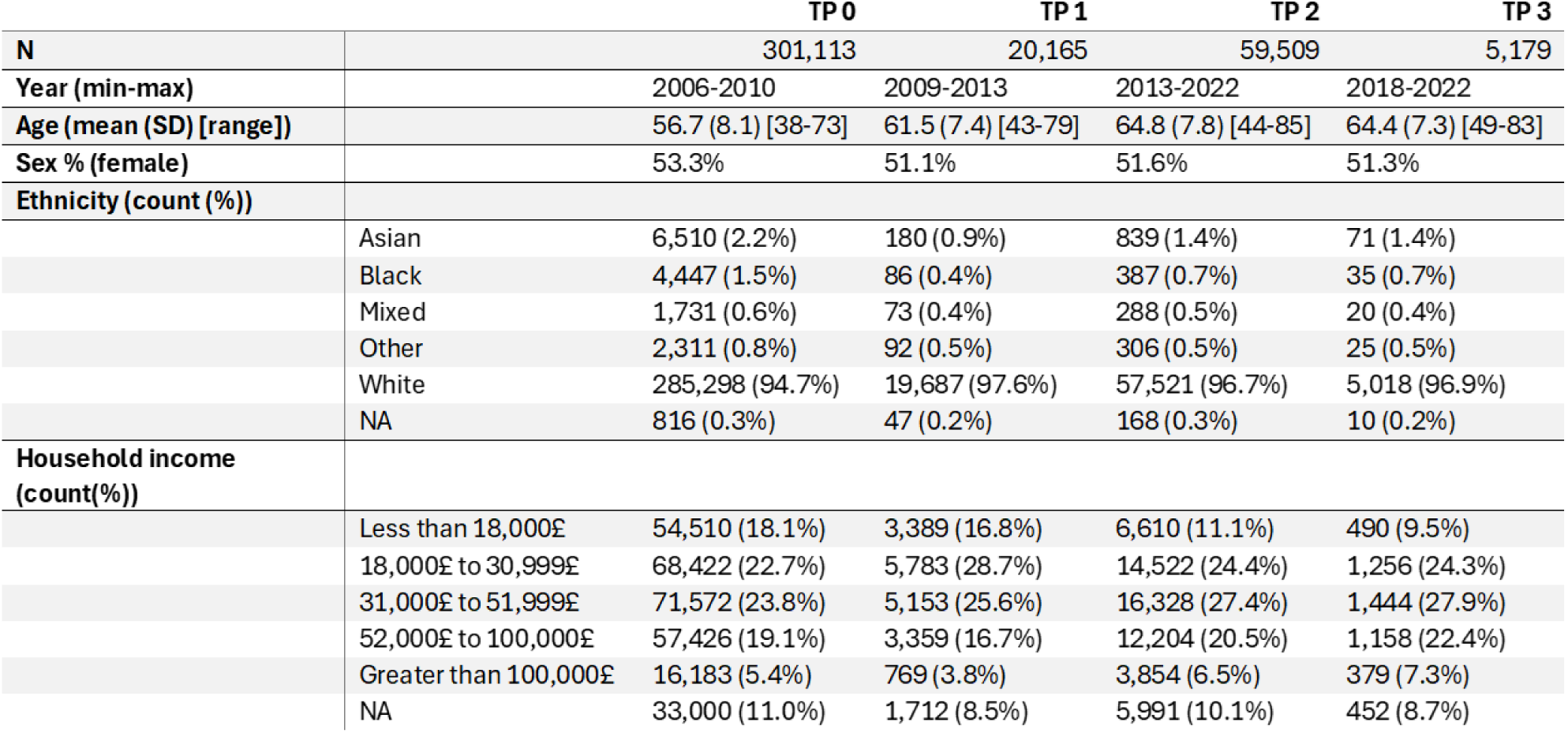
Demographic characteristics of the UK Biobank samples across four timepoints. . Ethnicity was self-reported and categorized. Household income was self-reported and refers to the average total household income before tax. TP: timepoint; SD: standard deviation; NA: missing values

Our classifier achieved high accuracy (area under the receiver operating characteristic curve [AUC]=0.929; Fig. 1B), indicating substantial gender-related signal in the data. 15% of participants were classified with an IGL incongruent with their sex (Fig. 1C), reflecting meaningful within-sex variance in gendered behaviors (Fig. 1D). We used SHapley Additive exPlanations (SHAP) values to characterize the reconstructed gendered dynamics at the population, variable, and individual levels (Methods). Individual-level SHAP-based explanations are shown in Supp. Figs. 3–5, demonstrating the mosaic of masculine and feminine behaviors enacted by each person.

**Figure 3.**
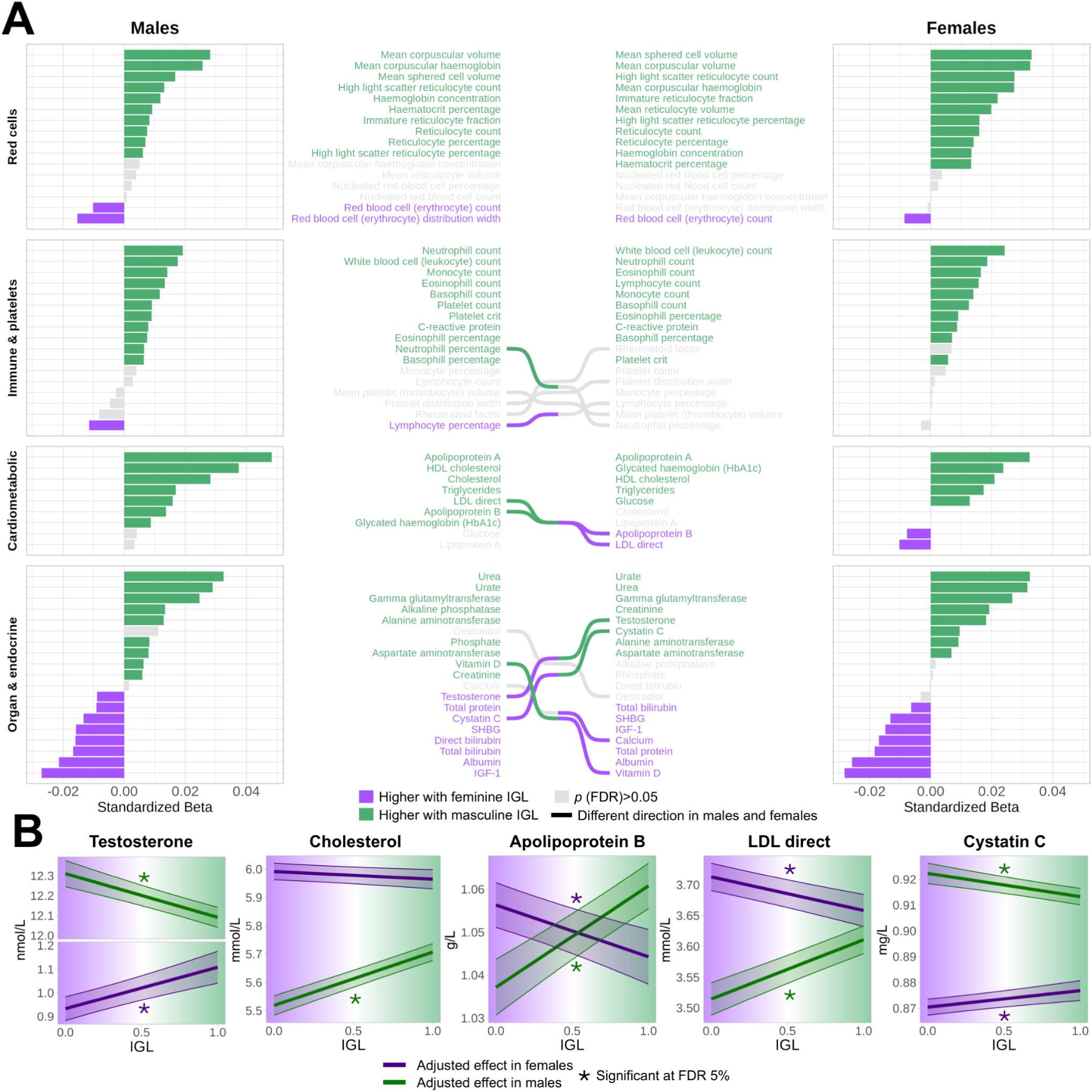
Associations between the IGL and blood biomarkers. **A)** Bar graphs represent the sex-specific standardized betas between the IGL and each blood biomarker after covariate adjustment. Positive effects indicate associations with a masculine IGL (light green) and negative effects indicate associations with a feminine IGL (light purple). Lines connecting markers across the bar graphs indicate an opposite association direction between males and females. **B)** Examples of individual associations that showed opposite effects in males and females. Fits represent effects after covariate adjustment and are color-coded according to sex (dark green for males and dark purple for females). Low-density lipoprotein (LDL), high-density lipoprotein (HDL), insulin-like growth factor 1 (IGF-1), sex hormone-binding globulin (SHBG).

**Figure 4.**
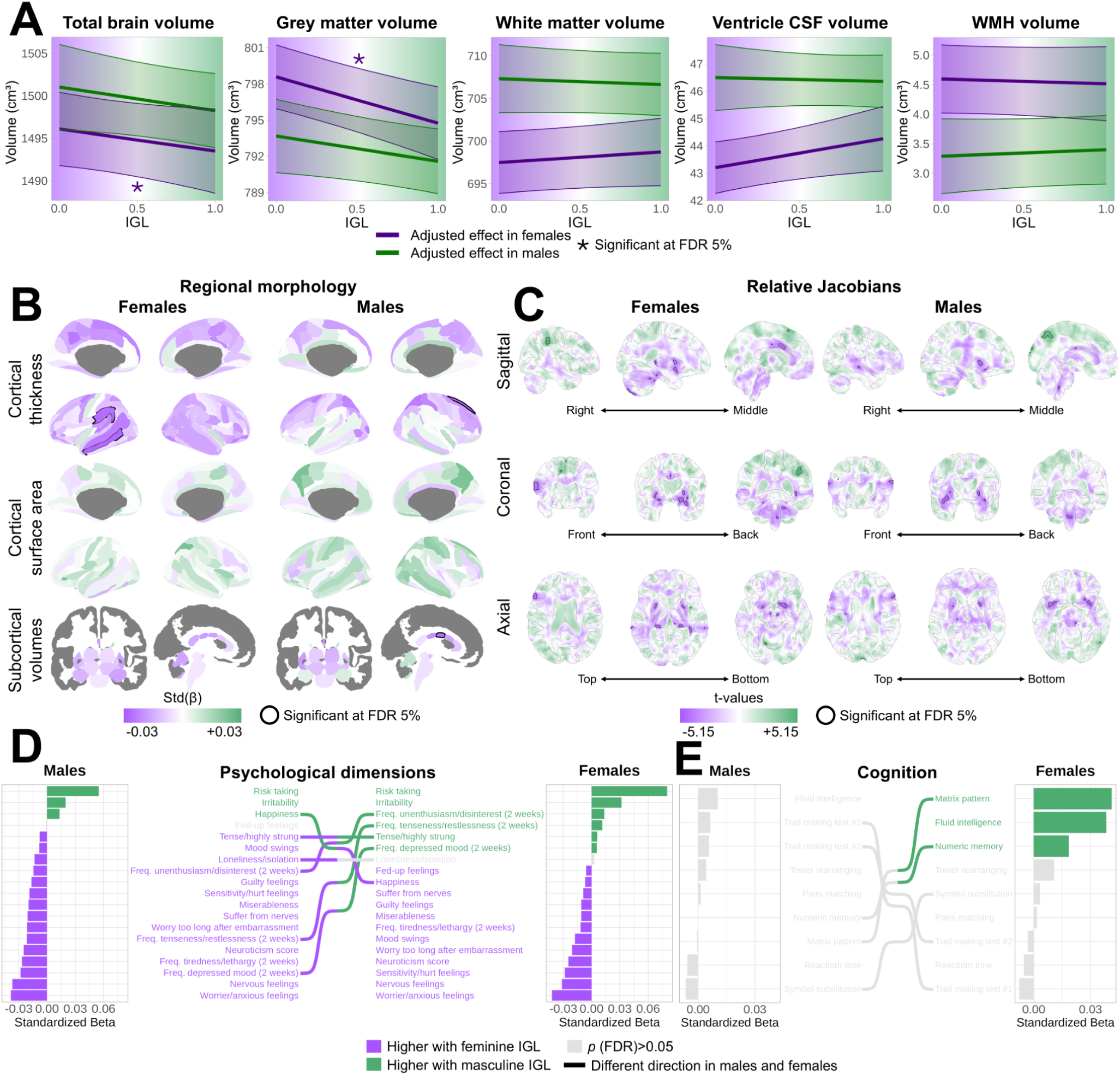
Associations between the IGL and brain morphology, psychological dimensions, and cognition. **A)** Associations between the IGL and whole-brain measures normalized for head size. Fits represent effects after covariate adjustment and are color-coded according to sex (dark green for males and dark purple for females). Associations between the IGL and **B)** region-of-interest measures of regional brain morphology (cortical thickness, cortical surface area, and subcortical structure volumes), and **C)** voxel-wise volumetric measurements (relative Jacobians) derived from deformation-based morphometry. Significant associations at the FDR 5% level are outlined in black. IGL associations with **D)** psychological dimensions and **E)** cognition are shown with bar graphs representing standardized betas calculated after covariate adjustment. Lines connecting markers across the bar graphs indicate an opposite association direction between males and females. Cerebrospinal fluid (CSF), white matter hyperintensity (WMH), false discovery rate (FDR), standardized beta (std(β)).

**Figure 5.**
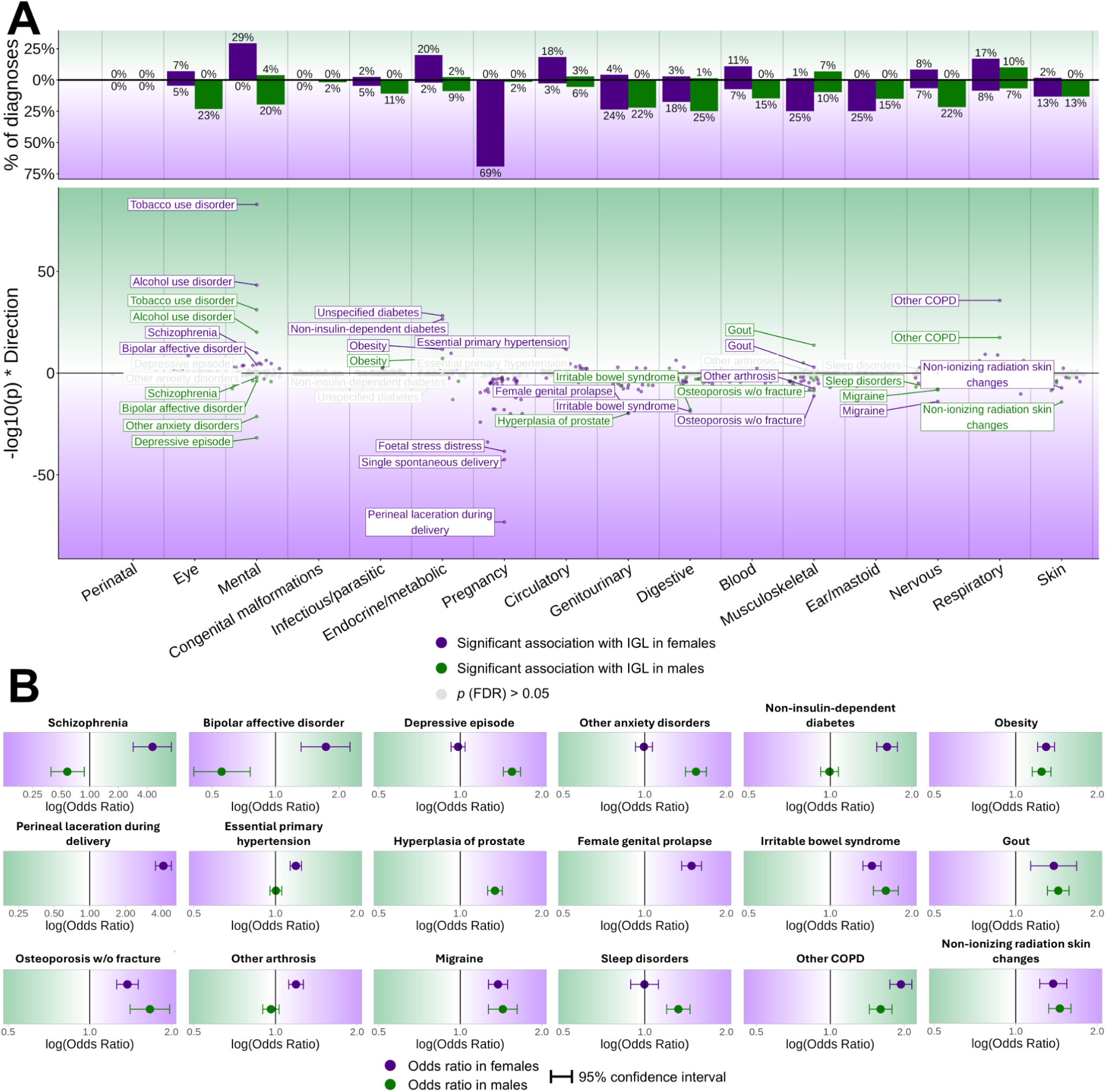
Associations between the IGL and medical diagnoses. Using medical records and ICD-10 codes, we associated the IGL with medical diagnoses in males and females separately using logistic regression. Control groups were defined as individuals without any diagnoses in the ICD-10 chapter of the diagnosis of interest. Significant effects at the FDR 5% level are shown. **A)** Top: the percentage of diagnoses showing significant effects in males and females (separately in the masculine and feminine direction) was calculated relative to the total number of diagnoses in each ICD-10 chapter. Bottom: Miami plot of significant effects in males (dark green) and females (dark purple). The y-axis represents -log_10_(*p*-values), with the feminine IGL direction indicated with negative values. Non-significant effects are shown in gray. Notable diagnosis effects are labeled. **B)** Examples of the effects of IGL on diagnosis prevalence, presented as sex-specific odds ratios with 95% confidence intervals relative to when the IGL is most masculine (IGL of 1) or most feminine (IGL of 0).

Population-typical masculine and feminine behaviors spanned all four domains (Fig. 1E–F). Masculine predictors clustered around substance use, work-related behaviors, calorie-dense diets, and high-impact physical activities. Feminine predictors included health-conscious practices (skin care, sleep routines), social activities, having a caretaker role, and low-impact physical activities. Several effects were non-linear (Supp. Figs. 6–9), with mobile phone adoption showing a U-shaped gender effect and work week length showing a threshold effect (<38 hours being more feminine and <38 hours masculine).

Although the IGL was not designed to measure gender identity, we hypothesized that it captured latent gender dimensions relevant to sexual and gender minorities. Both self-reported females with male chromosomes (*n*=71) and self-reported males with female chromosomes (*n*=29) without sex aneuploidies had, on average, more androgynous IGLs (i.e., less aligned with either gender profile) than sex-congruent individuals (*p*<0.01; Supp. Fig. 10). Similarly, both males and females who reported ever having had same-sex intercourse had more androgynous IGLs than participants who did not (*p*<0.0001). These preliminary findings suggest that gender and sexual minority individuals exhibit behavioral patterns less aligned with the dominant gender roles.

These gendered dynamics represent a snapshot of the United Kingdom circa 2006–2010. To investigate their evolution over the study period, we constructed IGLs at the three other timepoints (up to 2022), isolating changing gendered norms from shifting demographics by matching timepoints on age and ethnicity (Methods; Ext. Fig. 1), resulting in identical sample sizes (*n*=3,168). Several behaviors became less gendered over time, such as health behaviors (skin care, substance use), technology adoption, and educational attainment. On the other hand, some social behaviors became more gendered (age at first intercourse, group activities, family and friendship satisfaction), highlighting that gendered norms have not evolved uniformly across domains.

### Gendered dynamics across demographic dimensions

An important consideration in interpreting our results from the complete UKBB cohort is that participants were predominantly White (∼95%), spanned various socioeconomic strata, and covered a broad age range (38–73 years; Table 1). These demographic dimensions represent crucial sociocultural contexts shaping gendered norms and expectations,^15–17^ which may impact how gender associates with health phenotypes in different communities. We therefore characterized the contents of gendered dynamics across ethnicities, generations, and household income levels, matching each group on the other dimensions (Methods).

Individuals self-reported their ethnicities, categorized as White, Asian, and Black. Large demographic imbalances across unmatched groups (Supp. Fig. 11) were eliminated after matching (*n*=3,106 per group; Fig. 2A). The Asian category contained more South Asian than East Asian participants, while the Black category was more evenly split between African and Caribbean participants. Reconstruction accuracy was not significantly different between ethnicities, indicating similar overall gender effects (Fig. 2B), though gendered weightings differed substantially (Fig. 2C). In White participants, masculine effects were strongest for work week length and feminine effects for social visits, relative to other ethnic groups. In Asian participants, masculine effects were strongest for substance use and university degrees, and feminine effects for skin care. In Black participants, masculine effects were strongest for beef consumption and number of sexual partners, and feminine effects for high-school and professional degrees.

We also compared gendered dynamics within ethnic categories. A comparison of South Asian and Chinese participants showed that skin care was particularly feminine and smoking particularly masculine in Chinese participants (Supp. Fig. 12). Comparing African and Caribbean individuals revealed particularly strong feminine effects of having a professional degree in African participants (Supp. Fig. 13).

Across generations (*n*=18,046; Ext. Fig. 2), accuracy was higher in younger cohorts (AUC from 0.928 to 0.912), though this likely reflects younger generations being of working age rather than stronger gendered influences per se, given that work week length was among the few variables more gendered in younger generations. In contrast, many behaviors were progressively less gendered in younger generations, including certain dietary choices, technology use, sleeping habits, substance use, social activities, and higher educational attainment. Of note, these effects represent a mix of societal shifts and age-related temporal trends.

Gendered dynamics across household income (*n*=12,430; Ext. Fig. 3) were most rigidly differentiated in middle-income households (AUC=0.927, versus 0.902 and 0.914 in lower- and higher-income households) and converged on specific labor effects, with higher masculine effects for higher education and computer use. In lower-income households, gendered behaviors centered on physical labor, with masculine effects strongest for disability leave and time spent outdoors/driving. In higher-income households, gendered dynamics showed a marked division between masculine paid work and feminine domestic caregiving, with access to employment and education being largely ungendered.

Taken together, these findings demonstrate that gendered dynamics are not uniform across sociocultural contexts but are systematically shaped by ethnicity, generation, and socioeconomic position.

### Gender and body health

The IGL summarizes each participant’s proximity to population-typical masculine and feminine behavioral profiles into a single score that can be related to population health outcomes in males and females separately. This crucially allows gender to be investigated as a determinant of health beyond binary sex. We analyzed associations with body health markers using sex-stratified linear mixed-effects models with appropriate covariates such as age, assessment center, and body size (Methods; Supp. Table 2).

Blood-based biomarkers revealed numerous small (standardized β between 0.01 and 0.04) but significant associations across biomarker domains (Fig. 3A). In males and females, a masculine IGL was collectively associated with biochemistry consistent with elevated metabolic load and hepatic processing demands (higher urate, urea, and gamma glutamyltransferase; lower albumin), while a feminine IGL was associated with a growth-promoting anabolic state (higher circulating insulin-like growth factor 1 [IGF-1]). Several lipid and hormonal biomarkers exhibited opposite associations with the IGL in males and females, demonstrating interactions between gendered behaviors and male/female physiology (Fig. 3B). Testosterone was lower in masculine males but higher in masculine females. This could reflect the negative impact of substance use on testosterone,^18^ and the positive bidirectional effects between testosterone and masculine behaviors,^19,20^ particularly in females. Similarly, lipid markers (cholesterol, apolipoprotein B, low-density lipoprotein [LDL] direct) showed positive associations with masculinity for males but negative or null associations for females, which could be linked to estrogen-mediated lipid metabolism in females.^21^

Abdominal magnetic resonance imaging (MRI) data revealed anatomical associations corroborating these biochemical patterns. A masculine IGL was associated with larger organ volumes (liver, kidney, lung), higher liver iron, and greater cardiac left ventricle mass and wall thickness in both males and females (Ext. Figs. 4–5). Body composition measured with dual-energy X-ray absorptiometry (DXA) showed expected gendered correlates: a feminine IGL was associated with higher fat mass across the body, while a masculine IGL was associated with higher lean and bone mass (Ext. Fig. 6), consistent with the masculine behavioral profile of vigorous activity and calorie-dense diets.

Functional measures reflected these biochemical and structural patterns (Ext. Fig. 7). In line with DXA results, we observed that a masculine IGL was related to higher hand grip strength. A clear cardiovascular phenotype emerged, with a masculine IGL associated with higher blood pressure, arterial pressure and stiffness, and carotid intima-media thickness in males and females. A feminine IGL was associated with a higher heart rate during exercise and higher lung function, possibly reflecting feminine behaviors such as less strenuous physical activity and lower tobacco consumption.

Across biochemical, structural, and functional measures, gendered phenotypes were largely shared between males and females and demonstrated that gendered behavioral patterns capture physiological variance not explained by binary sex alone, although with generally small effect sizes (β∼0.01–0.05). A masculine IGL was broadly consistent with higher physiological throughput characterized by higher metabolic turnover, organ workload, and musculoskeletal demand, while a feminine IGL reflected lower throughput marked by greater energy storage and reduced systemic demand.

### Gender and brain health

We next asked whether gendered life patterns were reflected in markers of brain health. Global brain measures normalized for head size revealed female-specific associations, with a masculine IGL subtly related to lower total brain volume (β=−0.012; Fig. 4A), driven by lower gray matter (β=−0.024). These effects did not reach significance in males. The IGL was not significantly related to white matter hyperintensity (WMH) volume, which is sensitive to cerebral small vessel disease.^22^

To localize these effects, we investigated region-of-interest measures of cortical thickness, surface area, and subcortical volumes (Fig. 4B). A feminine IGL was associated with higher cortical thickness in left-hemisphere associative regions in females (middle temporal gyrus, parietal supramarginal gyrus), with a significant male effect isolated to the superior frontal sulcus. No significant effects emerged for surface area. A feminine IGL was associated with a larger middle corpus callosum in males, an isolated finding requiring replication. Investigating finer-grained brain anatomy (Methods; Fig. 4C; Supp. Methods 2; Supp. Fig. 14) revealed that a feminine IGL was associated with larger volumes in sensory and limbic regions in females (lateral olfactory gyrus, cerebellum, thalamic pulvinar, basal forebrain, nucleus accumbens) and in the putamen and thalamus in males. A masculine IGL was associated with larger volume in the language-processing supramarginal gyrus in females. Collectively, these findings indicate that a feminine IGL is associated with subtle (β∼0.01–0.03) structural effects in cerebral systems supporting social, emotional, and integrative processing, especially in females.

We next tested whether these structural findings were consistent with psychological and cognitive patterns. Gendered associations with personality traits were consistent across sexes and reproduced known sex differences,^23,24^ such that a masculine IGL was related to higher risk-taking and irritability, while a feminine IGL was related to higher neuroticism (Fig. 4D). Trait-like anxiety and depression were associated with a feminine IGL in males and females (e.g., worrier/anxious feelings, miserableness). Many state-like symptoms, such as feeling depressed in the last two weeks, were higher in individuals with an incongruent IGL relative to their sex (i.e., feminine males and masculine females), suggesting that deviation from gendered norms carries psychological costs distinct from those of gendered behaviors themselves. Cognitively, females with a masculine IGL performed significantly better on abstract reasoning and working memory tasks (Fig. 4E), possibly reflecting masculine-typed factors of higher educational attainment.^25^ No significant effects were detected in males.

Together, these findings reveal a coherent pattern in which a feminine IGL is associated with greater limbic and associative brain volume, higher trait anxiety and neuroticism, and lower cognitive performance on specific tasks in females, while state-like psychological symptoms are associated with sex–IGL incongruence rather than femininity per se. A masculine IGL was associated with male-typical personality traits in both sexes.

### Gender and medical diagnoses

While effects of gendered life domains on biological systems and psychological dimensions were small (β∼0.01–0.06) and do not imply healthy or unhealthy states at the individual level, they may confer heightened disease risk and relate to diagnosis prevalence at the population level, which can have large societal impacts. We performed a scoping analysis relating the IGL to the sex-specific prevalence of 854 diagnoses (Methods), observing 192 significant effects in females and 128 significant effects in males (Fig. 5A–B). Of note, medical diagnoses reflect not only underlying pathophysiology but also symptom perception, help-seeking behavior, and clinical recognition, which show clear gendered influences.^26^

Associations with mental health diagnoses partially mirrored our symptom-level findings. Schizophrenia and bipolar affective disorder were more prevalent in sex–IGL incongruent individuals. Depression and anxiety diagnoses were elevated only in males with a feminine IGL, despite symptom-level associations in both sexes, likely reflecting help-seeking behavior. Consistent with the masculine pattern of higher physiological load and substance use, a masculine IGL was associated with higher prevalence of respiratory diagnoses (e.g., chronic obstructive pulmonary disease [COPD]) and gout in both sexes, as well as circulatory (e.g., hypertension) and metabolic/endocrine (e.g., diabetes) diagnoses, particularly in females. Consistent with the feminine pattern of musculoskeletal vulnerability and higher trait-like anxiety, feminine IGL associations included osteoporosis/arthrosis and diagnoses in the nervous (e.g., migraine) and digestive (e.g., irritable bowel syndrome) systems across sexes. Pregnancy-related diagnoses showed a higher prevalence in females with a feminine IGL, even after correcting for the number of live births (Supp. Fig. 15), which could stem from greater antenatal and postnatal care utilization.

Across several diagnoses where feminine behaviors would be expected to be protective (e.g., skin care against radiation-related skin changes, sleep routines against sleep disorders), individuals with a feminine IGL nonetheless showed higher diagnosis prevalence, consistent with a pattern of higher healthcare engagement. This was supported by our finding that the use of most medications was higher in feminine individuals (Ext. Fig. 8). Together, these findings underscore that masculine and feminine gendered patterns have distinct medical correlates, with neither end of the gender spectrum being uniformly healthier than the other.

These disease risk profiles were derived from the cohort-wide IGL trained on a predominantly White sample and may not capture how gendered behaviors relate to health in ethnic minorities, particularly given the meaningful differences in behavioral gender weights we observed across ethnic groups. To address this, we constructed ethnicity-specific IGLs by applying models trained on matched White, Asian, and Black individuals to the complete sample, effectively reweighting each participant’s score according to the gendered norms of each ethnic sociocultural context, rather than analyzing biological ancestry (Methods).

When investigating associations between the Asian IGL and diagnosis prevalence (Ext. Fig. 9), we observed significantly different effects relative to the cohort-wide IGL in 190 diagnoses for males and 196 diagnoses for females. Strikingly, almost all of these interactions were in the masculine direction, indicating either a lower risk of a feminine IGL (e.g., irritable bowel syndrome), a higher risk of a masculine IGL (e.g., obesity, diabetes, hypertension, COPD), or even a reversal of risk from the feminine to the masculine direction (e.g., sleep disorders). This suggests that in Asian communities, where substance use and work-related behaviors are particularly strongly gendered, adherence to masculine norms carries a relatively stronger disease risk profile than in the broader cohort.

The Black IGL also showed differences in diagnosis prevalence relative to the cohort-wide IGL (Ext. Fig. 10), with 98 significant interactions in males and 58 significant interactions in females. Interestingly, we observed that pregnancy-related diagnoses showed higher prevalence in females with more masculine IGLs, although these results were heavily attenuated when controlling for number of live births (Supp. Fig. 16). Other significant interactions indicated increased masculine effects largely specific to males, including for circulatory (e.g., hypertension), musculoskeletal (e.g., gout), and respiratory (e.g., COPD) diagnoses. These findings highlight that a single population-wide gender score risks obscuring clinically meaningful heterogeneity across ethnic communities, arguing for sociocultural context as an essential parameter in gender-health research.

## Discussion

Epidemiological health research has long relied on binary sex as a proxy for the broader biopsychosocial influences that shape disease risk, neglecting the impact of socioculturally embedded gendered behaviors on health. Using population-typical behaviors more frequently expressed by males or females to define masculine and feminine behaviors, we developed the Index of Gendered Life-domains (IGL), a data-driven measure of how closely each individual aligns with prototypical gendered patterns. Within-sex variance in gendered behavior was systematically associated with health markers spanning blood biochemistry, organ physiology, brain morphology, psychological traits, and disease prevalence, establishing gender as a measurable and consequential determinant of health that can be used to monitor epidemiological trends in diverse communities.

Our investigation across biochemical, structural, and functional dimensions of body health showed how masculine behavioral patterns (e.g., vigorous activity, calorie-dense diets, substance use) were reflected in higher physiological load and elevated metabolic and circulatory disease risk, while feminine behavioral patterns were reflected in lower physiological throughput, reduced lean and bone mass, and vulnerability to musculoskeletal diseases. While risk factors for these diseases are well established,^27^ our study demonstrates that behaviors cluster in ways that reflect the biopsychosocial influences of gender, and that within each sex, conformity to gendered norms relates to disease risk profiles. This raises questions about how much of the male bias in cardiovascular and metabolic disease,^28,29^ and the female bias in musculoskeletal disease,^30^ reflects gender rather than sex alone, further highlighting the need for assessment of sex and gender simultaneously in epidemiological studies. Surprisingly, prior gender score studies reported that femininity was associated with worse cardiovascular outcomes,^13,31–35^ which may reflect their focus on psychosocial predictors^12^ capturing unhealthy aspects of femininity (e.g., anxiety and neuroticism) while missing those of masculinity (e.g., diet and substance use), a gap our approach addresses by focusing on health-related behaviors as core gendered predictors. Previous gender score implementations were also limited by small sample sizes and linearity assumptions.^12,13,36^

Our findings relating gender to brain health revealed two distinct patterns. First, femininity was associated with neuroticism and trait-like depression and anxiety, accompanied by subtle structural differences in limbic and associative brain regions particularly in females, and reflected in higher rates of medically diagnosed anxiety and depression particularly in males. This male-specificity of diagnoses likely reflects well-established gendered patterns of feminine help-seeking behavior.^37,38^ We now demonstrate this at the population scale, highlighting that the IGL captures not only disease risk but also healthcare engagement patterns influencing administrative health records. Collectively, these links between femininity and anxiety/depression are consistent with other gender score-based studies^36,39–41^ and the well-documented psychological burden of feminine gender roles such as caregiving.^42^ Second, sex–IGL incongruence was associated with state-like anxiety and depression symptoms and psychiatric diagnoses like schizophrenia and bipolar affective disorder. These observations resonate with minority stress theory, whereby stigma and reduced social support elevate psychiatric risk among sexual and gender identity minorities.^5,43^ Our results suggest these effects may extend to cisgender individuals conforming less to gendered expectations. Alternatively, disorganized behavior typical of schizophrenia could reduce conformity to gender norms. Together, these effects demonstrate that gender associates with mental health both through femininity relating with trait-level vulnerability and behavioral gender nonconformity relating with state-level psychiatric risk.

Our findings provide compelling evidence that health outcomes depend not only on adherence to health behaviors but also on how those behaviors align with normative gender expectations. Furthermore, these findings on sex–IGL incongruence underscore that gender is not simply the sum of its individual behaviors but instead represents a latent behavioral dimension, motivating the measurement of data-driven gender scores alongside individual behaviors in epidemiological research.

Gendered behavioral dynamics do not relate to health uniformly across communities. Rather than assuming masculine and feminine behaviors carry a fixed meaning, we adopted an intercategorical approach from intersectionality theory^44^ by mapping how the content of gendered behaviors is organized differently across ethnicities. The IGL approach allowed for both direct comparisons of gendered content between communities and prediction-based reweighting of gendered behaviors to empirically investigate gender-health associations in minority groups with high statistical power. This revealed that adherence to masculine norms in Asian communities, where substance use is particularly strongly gendered, was associated with substantially worse health outcomes than adherence to cohort-wide masculine norms, consistent with elevated cardiometabolic risk in South Asian populations^45^ that we frame through the lens of gendered behavioral clustering. Similarly, the relationship between feminine norms and pregnancy-related diagnoses showed a markedly opposite pattern in Black communities, consistent with documented structural and qualitative differences in how Black women in the UK access and experience maternity care.^46,47^ These findings argue that context-sensitive gender measurement is paramount for accurate health research in diverse populations. Accordingly, gender-health relationships derived from predominantly White samples risk misrepresenting the gendered behavioral profiles of ethnic minorities by applying majority-group norms as a universal standard.

Our work has several limitations. First, collapsing gendered factors into a single score obscures heterogeneity in individual gendered behavioral loadings,^48^ which may be addressed in future studies with domain-specific IGL subscores and by exploring each participant’s SHAP-based behavioral contributions. Second, the gendered dynamics described here reflect the UKBB’s specific sociocultural context, which may not generalize to other populations. As gender varies across demographic contexts, reproducibility of the IGL approach, rather than exact replication of findings, should be the goal. We provide exact questionnaire items for all predictors to facilitate this (Supp. Methods 1). Third, our sociocultural analyses isolated demographic dimensions from each other and could not capture more complex intersectional interactions, which will require larger and more diverse datasets.^49^ Fourth, causal directions of gender-health associations remain to be established.

Our research argues that gender is not merely a demographic label but a measurable behavioral dimension that is associated with biological structure and function, shaped by sociocultural context, and irreducible to binary sex, establishing gender measurement as an essential component of population health research.

## Methods

### UK Biobank cohort

In this study, we used data from the UK Biobank (UKBB), a large population-based cohort with over 500,000 individuals in the total sample (application #45551). The study was approved by the North West Multicenter Research Ethics Committee (United Kingdom), and all participants provided written informed consent. Recruitment of individuals aged 40–69 years occurred between 2006 and 2010 for the initial time point. Residents of England, Scotland, and Wales were invited to various assessment centers, where they provided self-reported data about their health and lifestyles via computerized questionnaires and tests, completed on a touch screen interface.

Across four timepoints between 2006 and 2022 (see Table 1), blood samples were gathered, medical imaging data were acquired, and cognitive evaluations were conducted, resulting in a broad range of health phenotypes. Medical diagnoses were mapped to ICD-10 codes using data from self-reported medical conditions and medical record linkage (primary care, hospital inpatient, and death register) by UKBB. Participants were excluded only on the basis of data availability (described below), in order to retain a more representative sample of the population. Still, the UKBB sample has a small but meaningful healthy volunteer selection bias, with participants, for example, less likely to live in socioeconomically deprived areas and less likely to smoke or drink alcohol relative to the UK population.^50^ Sex was measured in two ways: 1) sex was recorded in the central UK registry, which participants had the option to update, and 2) chromosomal sex (i.e., the presence of X and Y chromosomes) was determined from genotyping analysis, which also identified individuals with sex chromosome aneuploidies (i.e., sex chromosome configurations other than XX or XY).

### Selection and preprocessing of gender-related predictors

Variables included as predictors when constructing the IGL were acquired at all four timepoints and were chosen to represent the categories of diet, lifestyle (skin care, technology use, substance use, sleeping habits, hobbies, physical activity), social (sexual relations, household composition, social activities, relationship satisfaction), and socioeconomic factors (work, education, access to services). Variable selection within these categories was guided by data availability and theoretical relevance rather than prior empirical associations with sex or gender, in order to reduce confirmation bias. We focused on behavioral and socioeconomic factors that are not fixed by biological sex, since these could instead be influenced by gender via biopsychosocial factors.

Variables were cleaned to make them suitable as inputs to our machine learning model (Supp. Table 1). This included one-hot encoding categorical variables and assigning numerical values for ordinal factors. Since machine learning algorithms require complete data, missing data were imputed in a three-step hierarchy designed so that the most certain imputations were performed first. First, logical imputation handled cases where the true value was derivable from other variables (e.g., if the participant never smoked, the pack-years smoking variable was imputed as 0). Second, longitudinal imputation leveraged repeated measurements to fill remaining gaps (first from earlier to later timepoints, and second from later to earlier timepoints). After this step, participants with more than 10% of missing data across all variables were excluded to balance sample size retention with data quality, with the final sample sizes and demographics presented in Table 1. Third, algorithmic multiple imputation through XGBoost using the mixgb package (v1.5.3) in R (v4.4.0) addressed residual missingness. This was performed separately for each timepoint with 5 imputations per timepoint. Following imputation, the data were clamped to remove impossible values (e.g., if the number of days walking per week was below 0 or above 7, these were clamped to 0 and 7, respectively).

### Health phenotype preprocessing and neuroimaging pipeline

Some health phenotypes required additional cleaning steps before analysis. Systolic and diastolic blood pressure data were averaged across two measurements. Medication use data were mapped to anatomical therapeutic chemical (ATC) codes. Mental health variables were dichotomous or ordinal and underwent rank-based inverse normal transformation prior to inclusion as dependent variables in linear mixed-effects models. This procedure has statistical properties that are well-behaved for large, comparatively simple designs such as ours^51^ and allowed us to obtain standardized beta coefficients to remain consistent with other phenotypic associations. The use of linear mixed-effects models for binary and ordinal outcomes at this sample size is further supported by the close convergence of linear and logistic/ordinal regression estimates in large samples.^52^

White matter hyperintensity volume and region-of-interest cortical thickness, surface area, and subcortical volumes are imaging-derived phenotypes (IDPs) calculated by UKBB, with processing methods detailed elsewhere.^53,54^ To perform quality control on brain MRI data, we visually inspected T1-weighted (T1w) and fluid-attenuated inversion recovery (FLAIR) images according to guidelines established by our group and excluded scans showing significant motion artifacts (https://github.com/CoBrALab/documentation/wiki/Motion-Quality-Control-(QC)-Manual). To obtain voxel-wise measures of brain volume via deformation-based morphometry, we processed the brain MRI data using a custom pipeline developed by our group to maximize registration accuracy,^22^ detailed in Supp. Methods 2.

### Machine learning derivation of the index of gendered life-domains

To construct the Index of Gendered Life-domains (IGL), we predicted the sex of participants with the set of cleaned gendered variables described above using a machine learning algorithm. Participants with incongruent self-reported and chromosomal sex (*n*=176), as well as participants with sex chromosome aneuploidies (*n*=313), were held out from the training data. As a result, chromosomal sex and self-reported sex were identical variables, making these different sex definitions interchangeable as a classification target. We controlled for demographic sex differences in the training sample by matching males and females on age and ethnicity with the MatchIt package (v4.7.2) in R using exact matching, ensuring that our models would not be confounded by these characteristics or biased by an unequal number of males and females. We did not match males and females on income in order to maintain the signal related to socioeconomic inequality.

We used the MLjar-Supervised package (v1.1.18) in Python (v3.10.13) to train the models, an AutoML framework that automatically performed all appropriate processing and training steps, including scaling and hyperparameter tuning. An initial comparison of different non-linear algorithms on the complete baseline sample revealed that CatBoost models^55^ were the most accurate (Supp. Fig. 1) and were therefore used for all IGL constructions. Model training was repeated for the 5 imputed datasets, using 5-fold cross-validation repeated 5 times, resulting in 125 total models and 25 out-of-fold predictions per participant (except for participants held out from training, who have 125 predicted values). The IGL represents the sex classification probability averaged across runs. This was computed at each of the four timepoints to link the IGL to timepoint-specific health phenotypes (Supp. Fig. 2).

We then used SHapley Additive exPlanations (SHAP) values to examine the inner workings of the algorithm and the reconstructed gendered dynamics. SHAP values quantify the individual- and variable-specific contribution to the prediction, with positive values indicating that the data point shifted the prediction towards the positive class (male) and negative values towards the negative class (female). They provide insights at the model level (overall importance of predictors), variable level (shape of associations), and individual level (why each participant was given a specific prediction). For all 125 models, SHAP values were calculated on out-of-fold predictions in probability space for intuitive interpretation using the shap package (v0.49.1) in Python. Variable importance was calculated as the mean absolute SHAP value, averaged across models. Linear directions (masculine or feminine) were calculated for visualization purposes by computing Pearson correlations between SHAP values and raw values, again averaged across runs.

To construct IGL models within specific ethnic, age, and socioeconomic subgroups, as well as across timepoints, we first used matching to isolate each demographic dimension from the others, with a specific framework designed to balance optimal matching and sample size maximization. For each analysis, we first matched the two subgroups that were the farthest apart (e.g., timepoints 0 and 3), using exact matching when necessary on dimensions that showed especially large deviations (e.g., age). Other subgroups (e.g., timepoints 1 and 2) were then matched to one of the previously matched subgroups (e.g., matched timepoint 0) using nearest-neighbor matching based on propensity scores calculated with logistic regression, thus resulting in identical sample sizes for all subgroups. AUC values were compared using unpaired DeLong tests with the pROC package (v1.19.1) in R. Birth year cut-off values for generations followed guidelines from the Beresford research group (https://www.beresfordresearch.com/age-range-by-generation/). IGL models were then constructed within these subsets as described above.

### Associations with health phenotypes

We tested sex-stratified associations between the IGL and various health phenotypes with either linear mixed-effects models or logistic regression, where appropriate, using the *lmer* and *glm* functions in R (v4.4). This approach importantly isolates the within-sex gendered variance and its health correlates from effects of sex differences. The exact models are shown in Supp. Table 2. To obtain standardized beta coefficients in linear mixed-effects models, both the IGL and the phenotype of interest were z-scored. Models included covariates of non-linear age, modeled with third-degree B-splines with 4 degrees of freedom, and a random effect of assessment center, with exceptions (Supp. Table 2). Within each analysis, *p*-values were corrected for multiple comparisons controlling the false discovery rate at 5%.^56^ Sample sizes varied based on the availability of health phenotype data, and participants with missing values for any covariate were excluded. Imaging and cognitive phenotypes were only measured at timepoint 2 and are therefore related to the IGL constructed at timepoint 2 on the unmatched complete sample (Supp. Fig. 2).

For body health phenotypes, body size measures were important confounds to remove in many cases, especially since they correlated with the IGL (height [male/female]: *r*=0.048/0.038; weight [male/female]: *r*=0.064/0.063). These were included as covariates where appropriate, which isolates gendered behavioral effects from size-mediated effects. Body size covariates were notably used for the DXA analysis, which allowed us to isolate relative body composition measurements independent of overall body size. Blood biomarker associations were additionally corrected for non-linear effects of day of year and time of day to remove the influence of seasonal and circadian variations. For the exercise electrocardiography (ECG) analyses, only participants who performed the test on the bicycle were included.

Brain MRI associations were additionally corrected for intracranial volume, an estimate of motion during the T1w acquisition, and scanner position. For our voxel-wise brain volume investigation, we analyzed relative Jacobian determinants from the described multispectral registration (Supp. Methods 2), which are calculated after linear scaling and are thus normalized for brain size.

Associations with medical diagnoses were tested with logistic regression predicting case–control status. Controls were defined as individuals with no recorded diagnoses in the same ICD-10 chapter as the diagnosis of interest in order to avoid confounding by shared disease mechanisms within chapters (e.g., for hypertension, controls had no lifetime circulatory diagnoses). We adjusted only for linear age effects, as modeling non-linear effects would risk overfitting, especially for diagnoses with a relatively lower number of cases. Odds ratios were calculated based on the unstandardized IGL and thus represent the risk when the IGL is most masculine (IGL of 1) or most feminine (IGL of 0). Diagnoses with fewer than 20 cases were excluded. For medication use, we applied a similar case–control framework with logistic regression, whereby controls were not using any medication in the ATC category of the medication of interest. This analysis also covered non-ATC medications, including common medications and supplements such as those for blood pressure and cholesterol, for which controls were simply participants not using the medication of interest.

To characterize how adherence to ethnicity-specific gender norms relates to health outcomes, IGL models trained on the matched ethnic groups described above were applied to the complete sample. Again, predicted probabilities across 125 models were averaged to obtain a single IGL value per participant. In essence, each ethnicity-specific model learns an internal representation of masculine and feminine behavioral prototypes as they exist within that sociocultural context. For example, a participant’s Asian IGL score reflects how closely their behaviors resemble the masculine or feminine prototype defined by Asian gendered norms, irrespective of whether they are themselves Asian. This process thus isolates ethnicity-specific gendered behaviors rather than biological ancestry while retaining the statistical power of the full sample to detect associations with medical outcomes. To identify diagnoses where ethnicity-specific gendered dynamics significantly differed from cohort-wide patterns, we tested an IGL-by-ethnicity interaction term, using the cohort-wide IGL as a reference, in models applied to the complete sample.

## Code availability

All analysis code is available on a public GitHub repository (https://github.com/CoBrALab/igl).

## Data Availability

Data from the UK Biobank are available to approved researchers via material transfer agreements (https://www.ukbiobank.ac.uk/enable-your-research/register).

## Acknowledgments

We wish to thank all UK Biobank participants and staff for their invaluable contributions. OP is funded by the Alzheimer Society of Canada and the Fonds de Recherche du Québec - Santé (FRQS). MMC receives salary support from the FRQS and from a James McGill Professorship. MMC also receives research support from Canadian Institutes of Health Research, Natural Sciences and Engineering Research Council – Canada, McGill University’s Healthy Brains, Healthy Lives (a Canada First Research Excellence Fund Initiative), Parkinson’s Society Canada, and the Douglas Research Centre. MD receives salary support from the FRQS and the Canada Research Chair Program, research support from Canadian Institutes of Health Research, Natural Sciences and Engineering Research Council – Canada, Alzheimer’s Society Research Program and Brain Canada. RPJ receives salary support from the FRQS (Jr1–269532: https://doi.org/10.69777/269532 & Jr2–332273: https://doi.org/10.69777/332273) and the Fondation de l’Institut universitaire en santé mentale de Montréal.

## Competing interests

The authors declare no competing interests.

## Extended Data

**Extended Figure 1.**
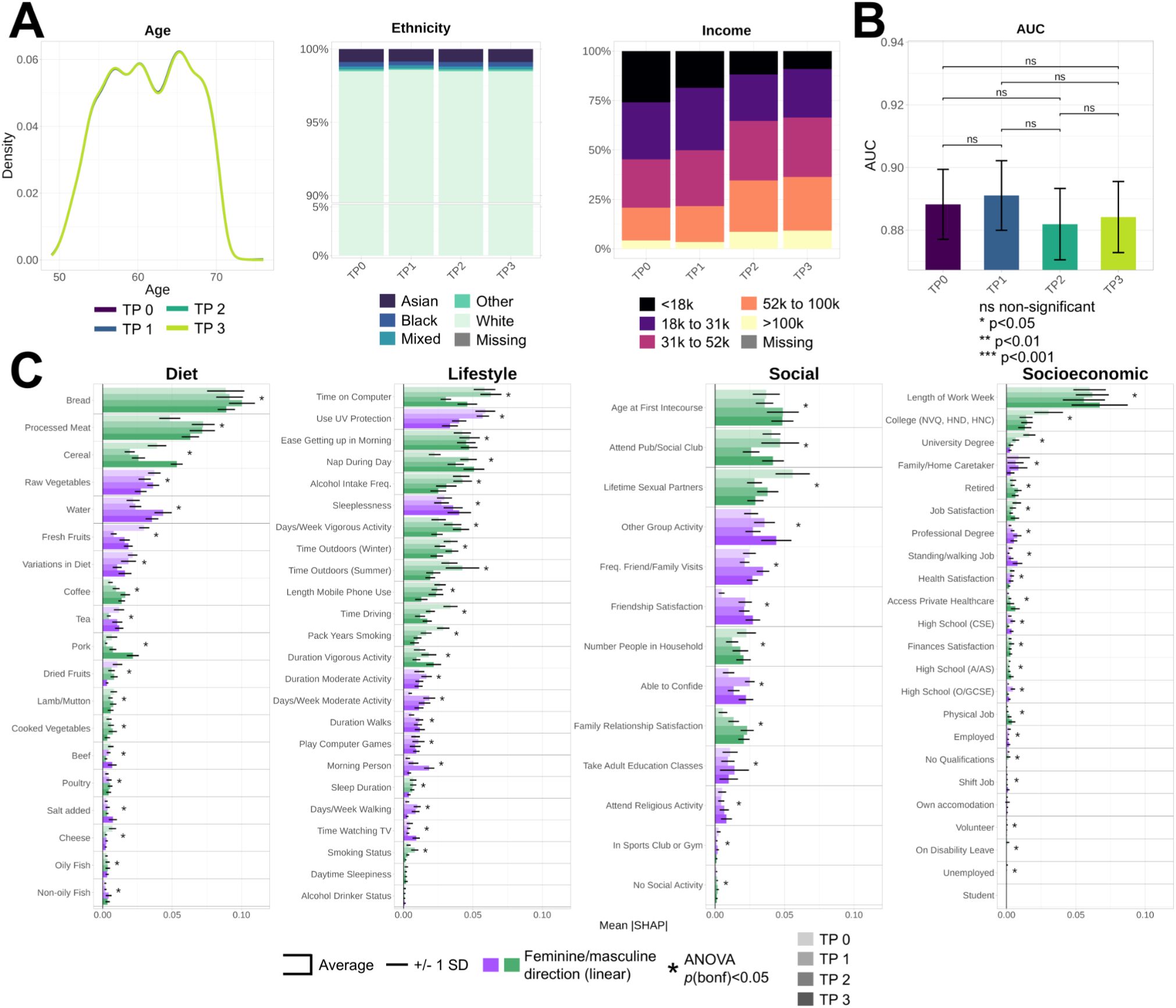
Gendered dynamics across timepoints. IGL models were computed at the four timepoints (TP) of the UKBB after matching groups on demographic variables. **A**) Distributions of age, ethnicity, and household income across timepoints. We observed almost perfect matching, with only timepoint one having marginally more White participants. **B**) AUC values were compared with unpaired DeLong tests. **C**) Mean absolute SHAP variable importances across timepoints (from light to dark). Error bars indicate +/- one standard deviation (SD). A linear direction was calculated with a Pearson correlation on the averaged SHAP plot and is indicated by a light purple (feminine) or light green (masculine) color. Statistical differences between TPs for each variable were calculated using ANOVAs, *p*-values were adjusted with a Bonferroni correction, and significant effects at *p*<0.05 are indicated with an asterisk.

**Extended Figure 2.**
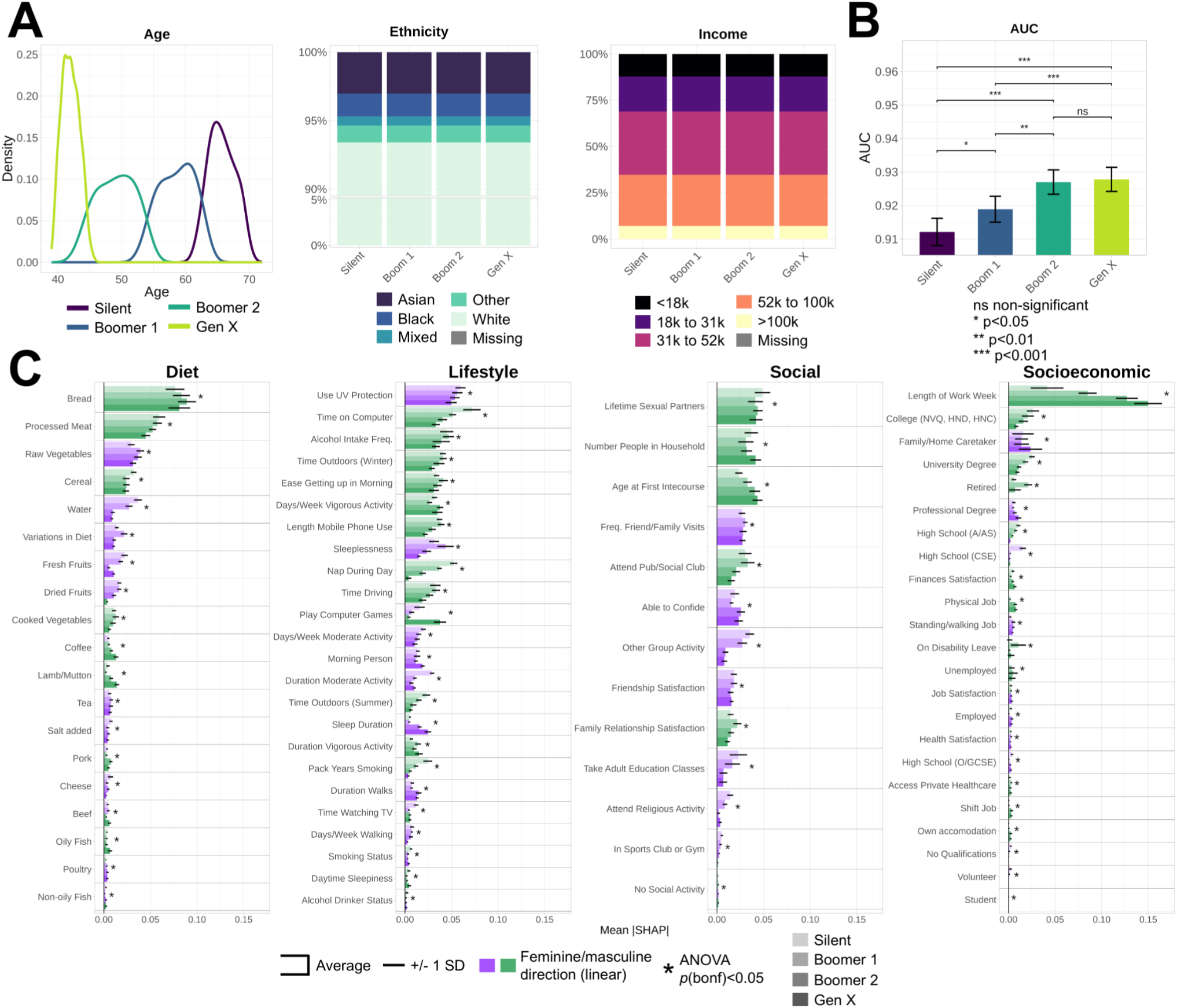
Gendered dynamics across generations. IGL models were computed in generation groups matched on ethnicity and household income. **A)** Distributions of age, ethnicity, and household income across generations. **B)** AUC values were compared with unpaired DeLong tests. **C)** Mean absolute SHAP variable importances across generations. Error bars indicate +/- one standard deviation (SD). For visualization purposes, a linear direction was calculated with a Pearson correlation on the averaged SHAP plot and is indicated by a light purple (feminine) or light green (masculine) color. Statistical differences between groups for each variable were calculated using ANOVAs, *p*-values were adjusted with a Bonferroni correction, and significant effects at *p*<0.05 are indicated with an asterisk.

**Extended Figure 3.**
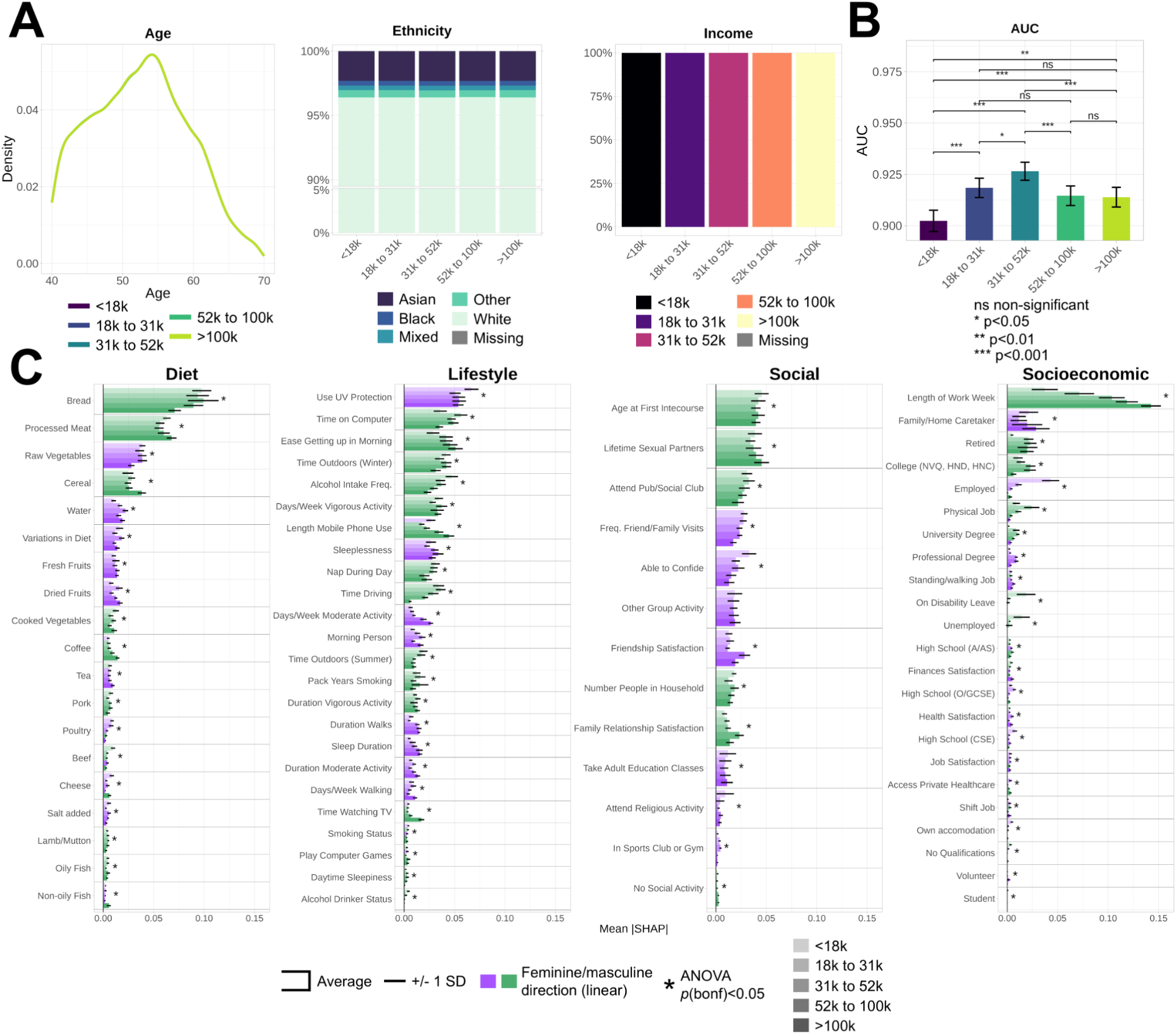
Gendered dynamics across income levels. IGL models were computed in household income groups matched on age and ethnicity. **A)** Distributions of age, ethnicity, and household income across income levels. **B)** AUC values were compared with unpaired DeLong tests. **C)** Mean absolute SHAP variable importances across income levels. Error bars indicate +/- one standard deviation (SD). For visualization purposes, a linear direction was calculated with a Pearson correlation on the averaged SHAP plot and is indicated by a light purple (feminine) or light green (masculine) color. Statistical differences between groups for each variable were calculated using ANOVAs, *p*-values were adjusted with a Bonferroni correction, and significant effects at *p*<0.05 are indicated with an asterisk.

**Extended Figure 4.**
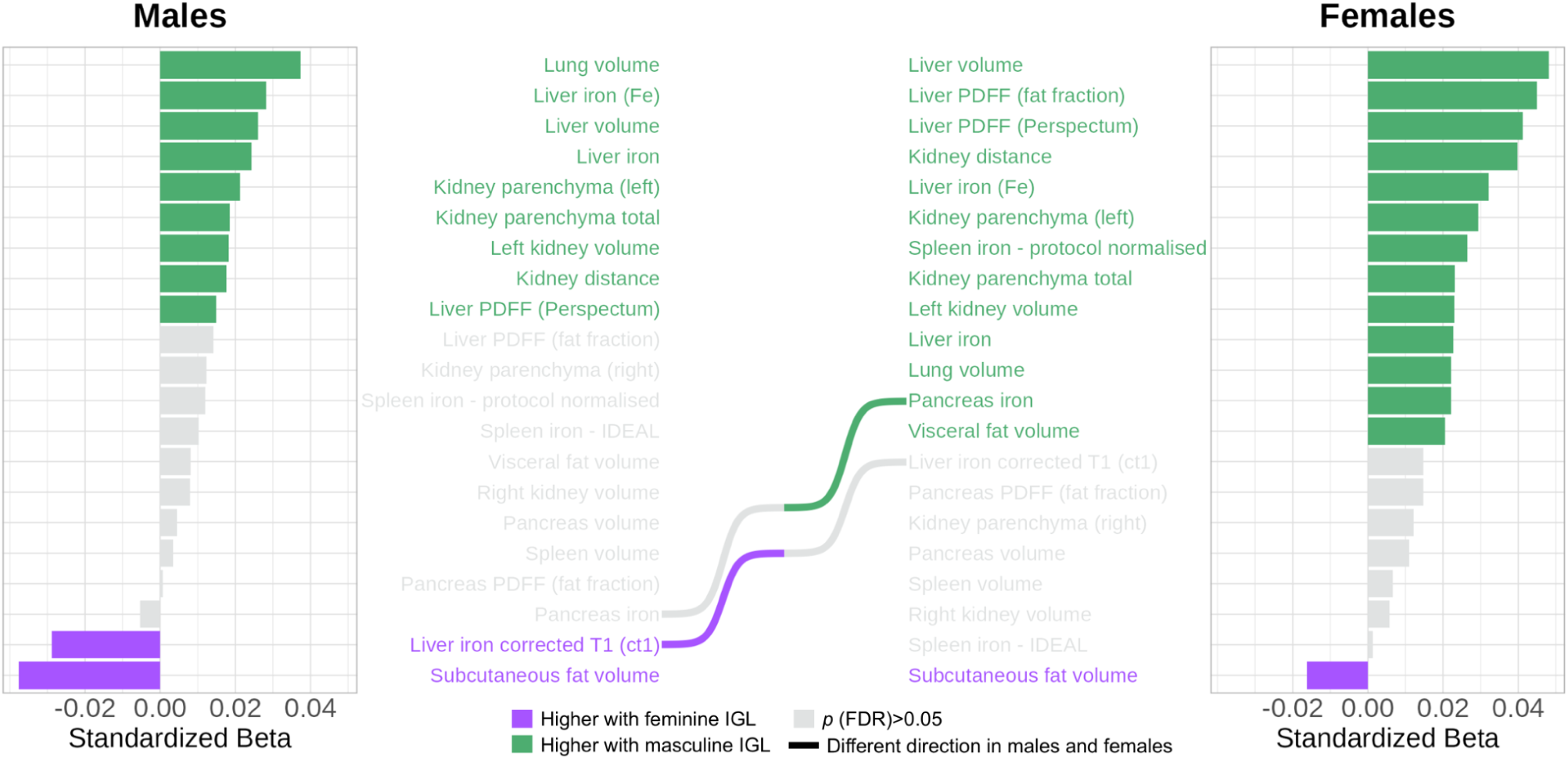
Associations with organ magnetic resonance imaging. Bar graphs represent the sex-specific standardized betas between the IGL and organ markers after covariate adjustment. Positive effects indicate associations with a masculine IGL (light green) and negative effects indicate associations with a feminine IGL (light purple). Lines connecting markers across the bar graphs indicate an opposite association direction between males and females.

**Extended Figure 5.**
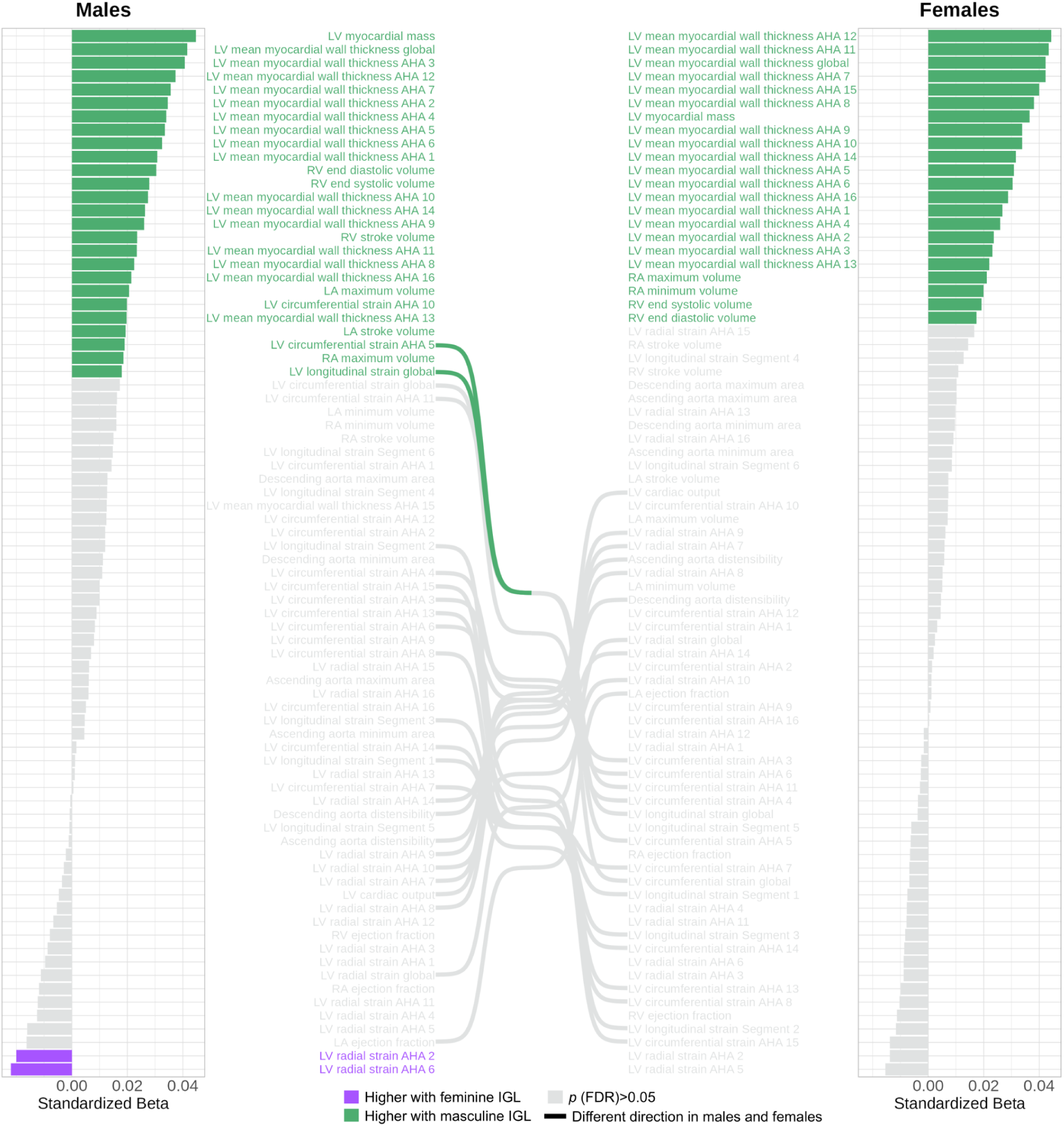
Associations with cardiac magnetic resonance imaging. Bar graphs represent the sex-specific standardized betas between the IGL and cardiac markers after covariate adjustment. Positive effects indicate associations with a masculine IGL (light green) and negative effects indicate associations with a feminine IGL (light purple). Lines connecting markers across the bar graphs indicate an opposite association direction between males and females.

**Extended Figure 6.**
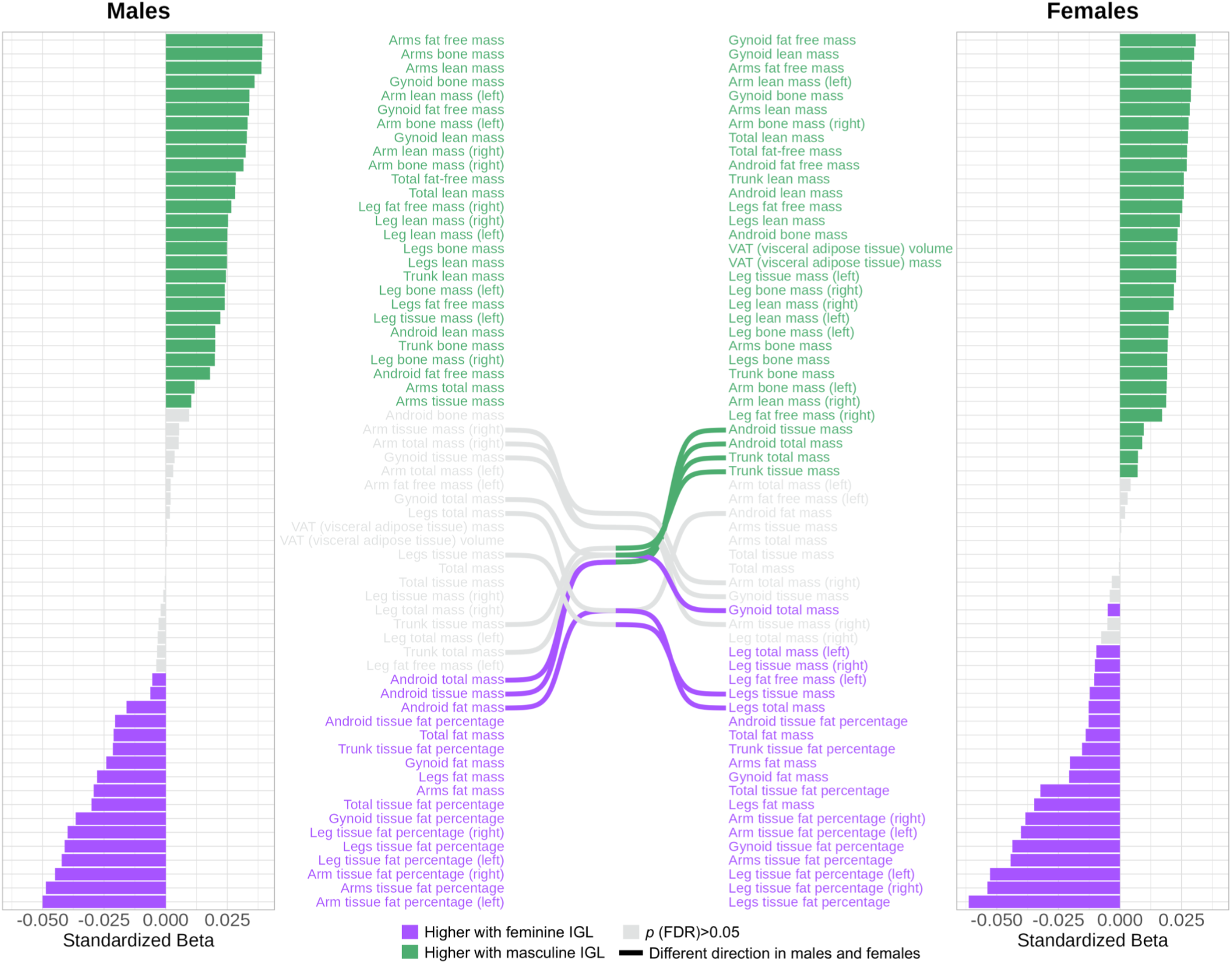
Associations with body composition from DXA measurements. Bar graphs represent the sex-specific standardized betas between the IGL and dual-energy X-ray absorptiometry (DXA) measurements after covariate adjustment. Of note, effects shown are corrected for height and weight and thus represent body composition relative to overall body size. Positive effects indicate associations with a masculine IGL (light green) and negative effects indicate associations with a feminine IGL (light purple). Lines connecting markers across the bar graphs indicate an opposite association direction between males and females.

**Extended Figure 7.**
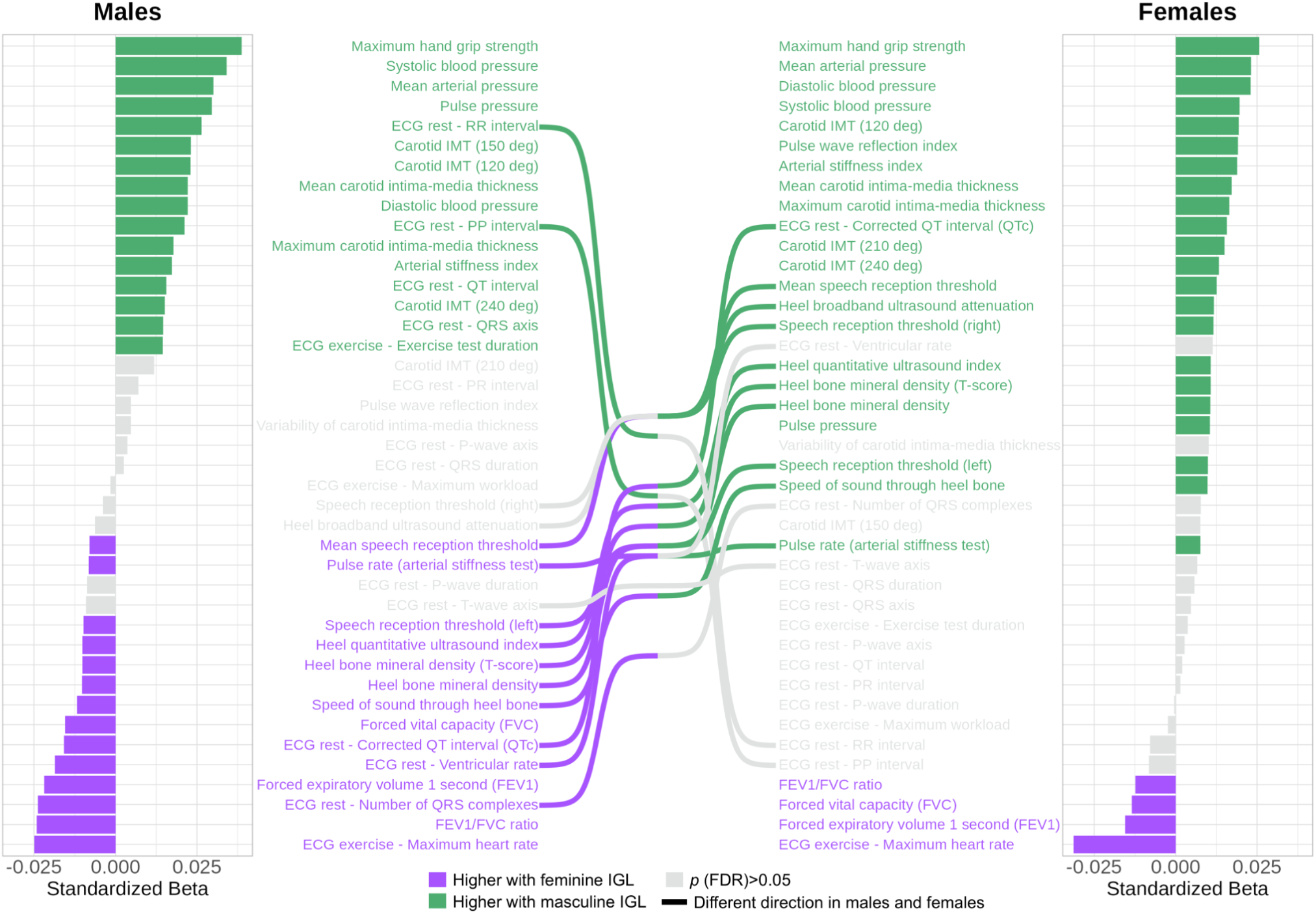
Associations with physical measures. Bar graphs represent the sex-specific standardized betas between the IGL and physical measures after covariate adjustment. Positive effects indicate associations with a masculine IGL (light green) and negative effects indicate associations with a feminine IGL (light purple). Lines connecting markers across the bar graphs indicate an opposite association direction between males and females.

**Extended Figure 8.**
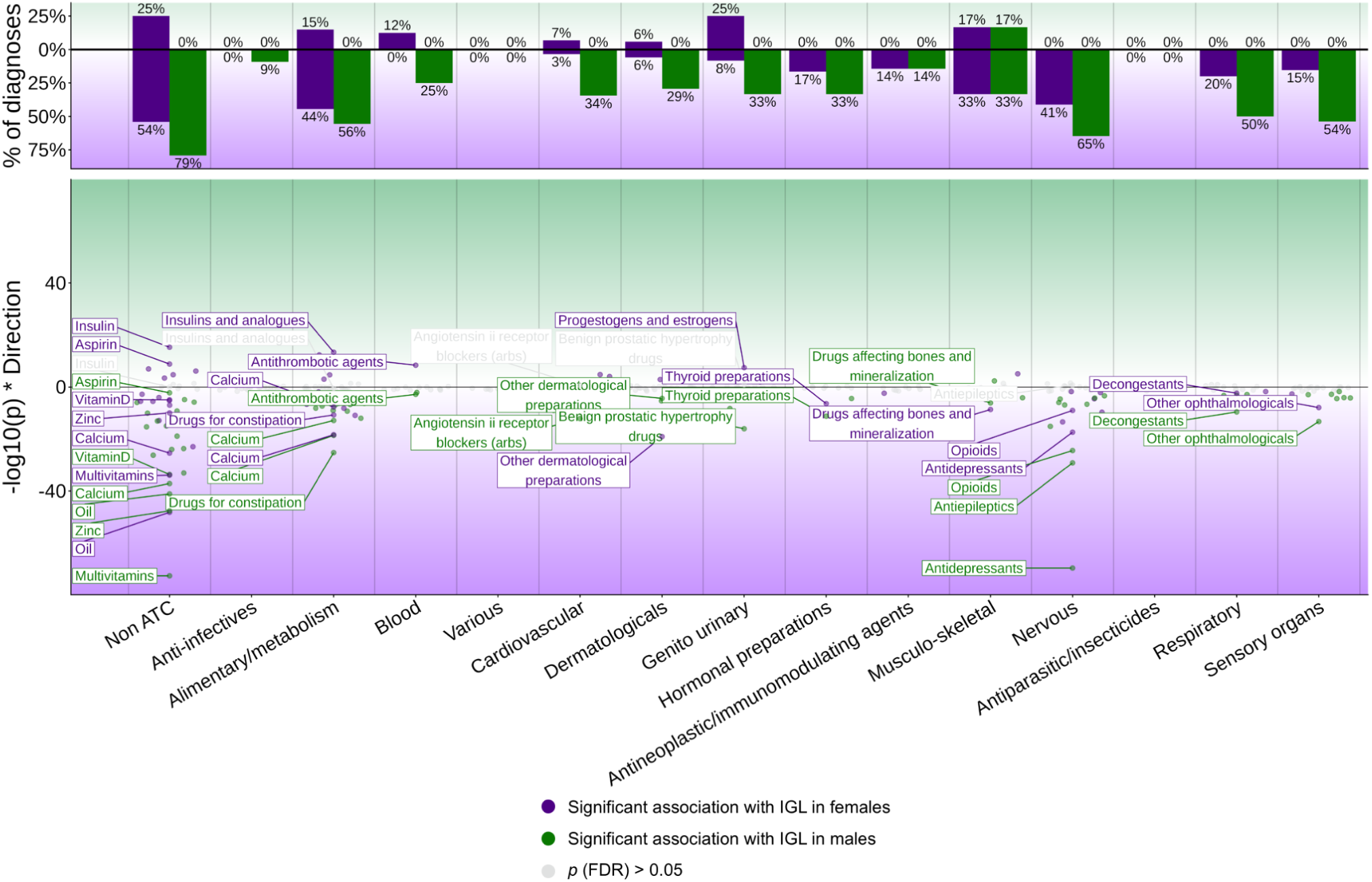
Associations with medication use. Using medication data mapped to anatomical therapeutic chemical (ATC) codes as well as non-ATC medications, we associated the IGL with medication use in males and females separately using logistic regression. Control groups were defined as individuals not using any medication in the ATC category of interest. Significant effects at the FDR 5% level are shown. Top: the percentage of medications showing significant effects in males and females (separately in the masculine and feminine direction) was calculated relative to the total number of medications in each category. Bottom: Miami plot of significant effects in males (dark green) and females (dark purple). The y-axis represents -log_10_(*p*-values), with the feminine IGL direction indicated with negative values. Non-significant effects are shown in gray. Notable medication effects are labeled.

**Extended Figure 9.**
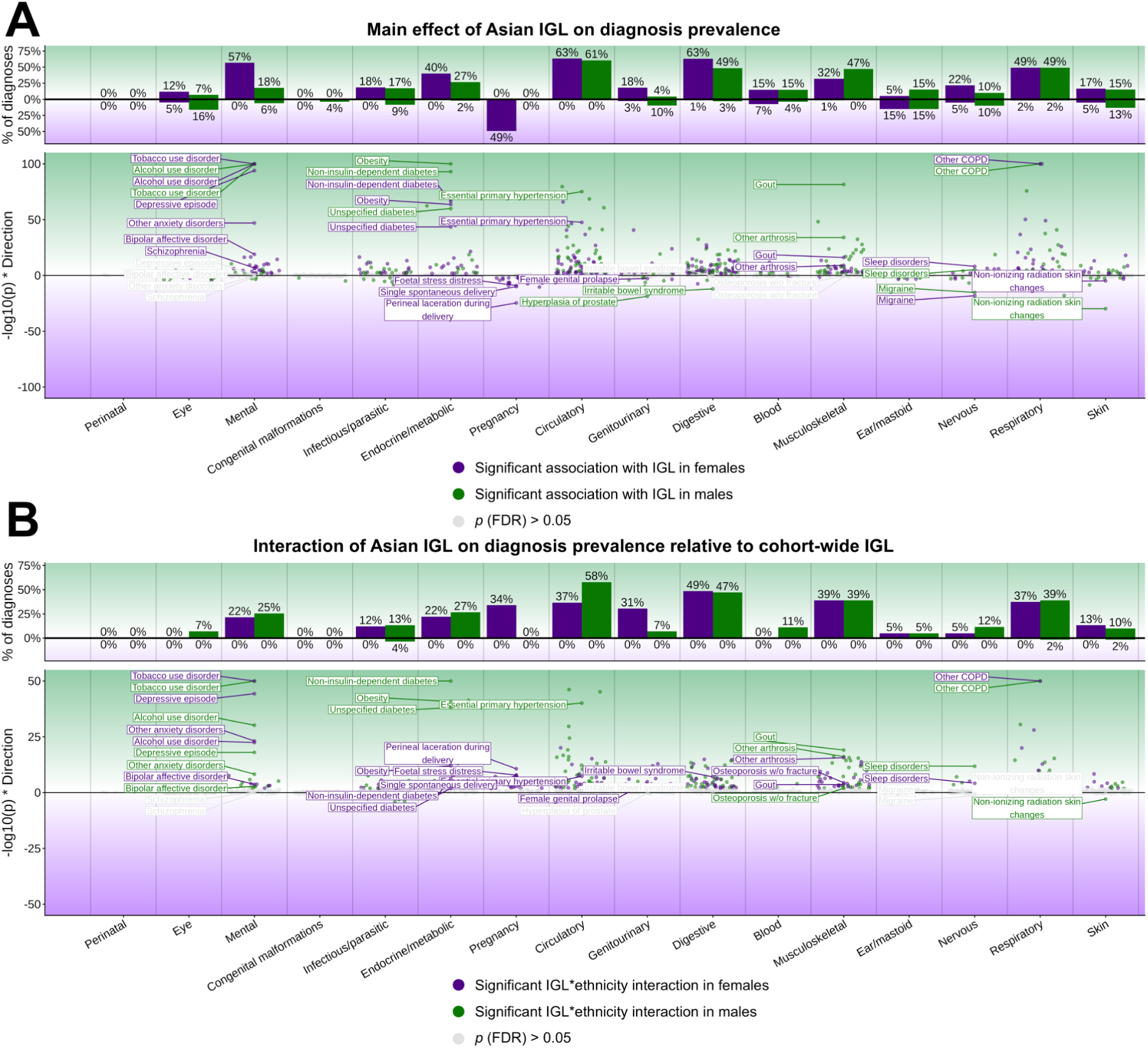
Associations between the Asian IGL and medical diagnoses. Using an IGL model constructed on matched Asian individuals, we related this IGL to medical diagnoses in males and females separately using logistic regression (**A**) and further assessed whether effects significantly differed from those calculated using the cohort-wide IGL with interaction terms (**B**). Control groups were defined as individuals without any diagnoses in the ICD-10 chapter of the diagnosis of interest. Significant effects at the FDR 5% level are shown. Top: the percentage of diagnoses showing significant effects in males and females (separately in the masculine and feminine direction) was calculated relative to the total number of diagnoses in each ICD-10 chapter. Bottom: Miami plot of significant effects in males (dark green) and females (dark purple). The y-axis represents -log_10_(*p*-values), with the feminine IGL direction indicated with negative values. Non-significant effects are shown in gray. Notable diagnosis effects are labeled.

**Extended Figure 10.**
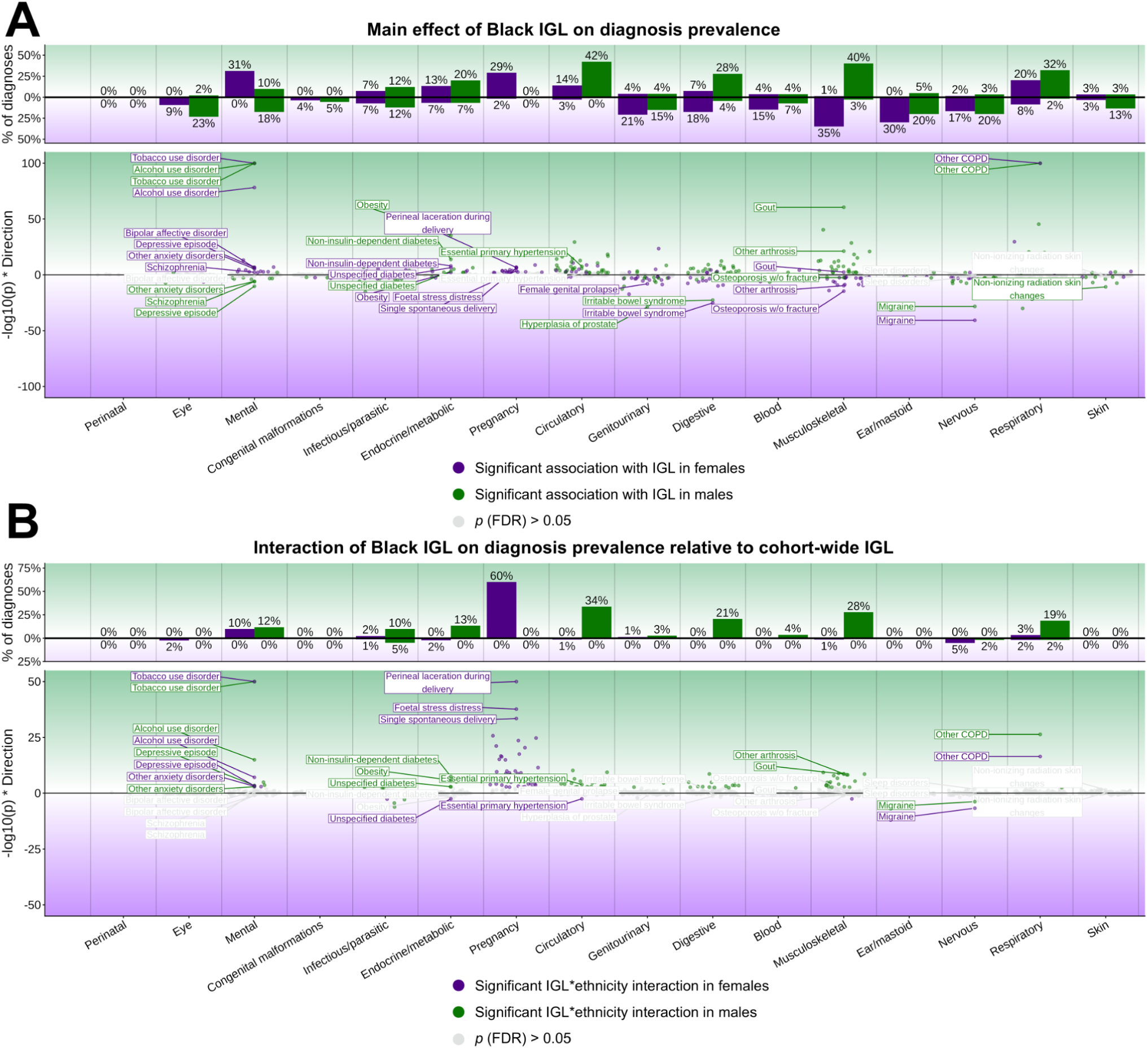
Associations between the Black IGL and medical diagnoses. Using an IGL model constructed on matched Black individuals, we related this IGL to medical diagnoses in males and females separately using logistic regression (**A**) and further assessed whether effects significantly differed from those calculated using the cohort-wide IGL with interaction terms (**B**). Control groups were defined as individuals without any diagnoses in the ICD-10 chapter of the diagnosis of interest. Significant effects at the FDR 5% level are shown. Top: the percentage of diagnoses showing significant effects in males and females (separately in the masculine and feminine direction) was calculated relative to the total number of diagnoses in each ICD-10 chapter. Bottom: Miami plot of significant effects in males (dark green) and females (dark purple). The y-axis represents -log_10_(*p*-values), with the feminine IGL direction indicated with negative values. Non-significant effects are shown in gray. Notable diagnosis effects are labeled.

## Notes

### Competing Interest Statement

The authors have declared no competing interest.

### Author Declarations

The study used only openly available human data that were originally located at https://www.ukbiobank.ac.uk/enable-your-research/register

