## Supplementary material for "Gendered behaviors are associated with health beyond binary sex": Supp.

**Supplementary Methods**.....3
Supplementary Methods 1. Questionnaire of gendered variables.....3
Supplementary Methods 2. Brain MRI processing for deformation-based morphometry. 10

### Supplementary Methods

#### Supplementary Methods 1. Questionnaire of gendered variables.

We list here the exact questions and answers for variables used as inputs to our gender score algorithm. These were administered on a touchscreen in the UK Biobank cohort. This information was taken from the UK Biobank Showcase browser (https://biobank.ndph.ox.ac.uk/showcase/).

**Diet**

- Cooked vegetable intake (fieldID 1289)
    - Question: On average how many heaped tablespoons of COOKED vegetables would you eat per DAY? (Do not include potatoes; put '0' if you do not eat any)
    - Choices: Numpad, less than one, do not know, prefer not to answer
- Salad / raw vegetable intake (fieldID 1299)
    - Question: On average how many heaped tablespoons of SALAD or RAW vegetables would you eat per DAY? (Include lettuce, tomato in sandwiches; put '0' if you do not eat any)
    - Choices: Numpad, less than one, do not know, prefer not to answer
- Fresh fruit intake (fieldID 1309)
    - Question: About how many pieces of FRESH fruit would you eat per DAY? (Count one apple, one banana, 10 grapes etc as one piece; put '0' if you do not eat any)
    - Choices: Numpad, less than one, do not know, prefer not to answer
- Dried fruit intake (fieldID 1319)
    - Question: About how many pieces of DRIED fruit would you eat per DAY? (Count one prune, one dried apricot, 10 raisins as one piece; put '0' if you do not eat any)
    - Choices: Numpad, less than one, do not know, prefer not to answer
- Oily fish intake (fieldID 1329)
    - Question: How often do you eat oily fish? (e.g. sardines, salmon, mackerel, herring).
    - Choices: Never, less than once a week, once a week, 2-4 times a week, 5-6 times a week, once or more daily, do not know, prefer not to answer.
- Non-oily fish intake (fieldID 1339)
    - Question: How often do you eat other types of fish? (e.g. cod, tinned tuna, haddock)
    - Choices: Never, less than once a week, once a week, 2-4 times a week, 5-6 times a week, once or more daily, do not know, prefer not to answer.
- Processed meat intake (fieldID 1349)
    - Question: How often do you eat processed meats (such as bacon, ham, sausages, meat pies, kebabs, burgers, chicken nuggets)?

    - Choices: Never, less than once a week, once a week, 2-4 times a week, 5-6 times a week, once or more daily, do not know, prefer not to answer.
- Poultry intake (fieldID 1359)
    - Question: How often do you eat chicken, turkey or other poultry? (Do not count processed meats)
    - Choices: Never, less than once a week, once a week, 2-4 times a week, 5-6 times a week, once or more daily, do not know, prefer not to answer.
- Beef intake (fieldID 1369)
    - Question: How often do you eat beef? (Do not count processed meats)
    - Choices: Never, less than once a week, once a week, 2-4 times a week, 5-6 times a week, once or more daily, do not know, prefer not to answer.
- Lamb/mutton intake (fieldID 1379)
    - Question: How often do you eat lamb/mutton? (Do not count processed meats)
    - Choices: Never, less than once a week, once a week, 2-4 times a week, 5-6 times a week, once or more daily, do not know, prefer not to answer.
- Pork intake (fieldID 1389)
    - Question: How often do you eat pork? (Do not count processed meats such as bacon or ham)
    - Choices: Never, less than once a week, once a week, 2-4 times a week, 5-6 times a week, once or more daily, do not know, prefer not to answer.
- Cheese intake (fieldID 1408)
    - Question: How often do you eat cheese? (Include cheese in pizzas, quiches, cheese sauce etc)
    - Choices: Never, less than once a week, once a week, 2-4 times a week, 5-6 times a week, once or more daily, do not know, prefer not to answer.
- Bread intake (fieldID 1438)
    - Question: How many slices of bread do you eat each WEEK?
    - Choices: Numpad, less than one, do not know, prefer not to answer
- Cereal intake (fieldID 1458)
    - Question: How many bowls of cereal do you eat a WEEK?
    - Choices: Numpad, less than one, do not know, prefer not to answer
- Salt added to food (fieldID 1478)
    - Question: Do you add salt to your food? (Do not include salt used in cooking)
    - Choices: Never/rarely, sometimes, usually, always, prefer not to answer
- Tea intake (fieldID 1488)
    - Question: How many cups of tea do you drink each DAY? (Include black and green tea)
    - Choices: Numpad, less than one, do not know, prefer not to answer
- Coffee intake (fieldID 1498)
    - Question: How many cups of coffee do you drink each DAY? (Include decaffeinated coffee)
    - Choices: Numpad, less than one, do not know, prefer not to answer
- Water intake (fieldID 1528)

    - Question: How many glasses of water do you drink each DAY?
    - Choices: Numpad, less than one, do not know, prefer not to answer
- Variation in diet (fieldID 1548)
    - Question: Does your diet vary much from week to week?
    - Choices: Never/rarely, sometimes, often, do not know, prefer not to answer

**Lifestyle**

- Duration of moderate activity (fieldID 894)
    - Question: How many minutes did you usually spend doing moderate activities on a typical DAY?
    - Choices: Numpad, do not know, prefer not to answer
- Duration of vigorous activity (fieldID 914)
    - Question: How many minutes did you usually spend doing vigorous activities on a typical DAY?
    - Choices: Numpad, do not know, prefer not to answer
- Duration of walks (fieldID 874)
    - Question: How many minutes did you usually spend walking on a typical DAY?
    - Choices: Numpad, do not know, prefer not to answer
- Number of days/week of moderate physical activity 10+ minutes (fieldID 884)
    - Question: In a typical WEEK, on how many days did you do 10 minutes or more of moderate physical activities like carrying light loads, cycling at normal pace? (Do not include walking)
    - Choices: Numpad, do not know, prefer not to answer
- Number of days/week of vigorous physical activity 10+ minutes (fieldID 904)
    - Question: In a typical WEEK, how many days did you do 10 minutes or more of vigorous physical activity? (These are activities that make you sweat or breathe hard such as fast cycling, aerobics, heavy lifting)
    - Choices: Numpad, do not know, prefer not to answer
- Number of days/week walked 10+ minutes (fieldID 864)
    - Question: In a typical WEEK, on how many days did you walk for at least 10 minutes at a time? (Include walking that you do at work, travelling to and from work, and for sport or leisure)
    - Choices: Numpad, do not know, unable to walk, prefer not to answer
- Time spent driving (fieldID 1090)
    - Question: In a typical DAY, how many hours do you spend driving?
    - Choices: Numpad, less than an hour a day, do not know, prefer not to answer
- Time spent using computer (fieldID 1080)
    - Question: In a typical DAY, how many hours do you spend using the computer? (Do not include using a computer at work; put 0 if you do not spend any time doing it)
    - Choices: Numpad, less than an hour a day, do not know, prefer not to answer
- Time spent watching television (TV) (fieldID 1070)

    - Question: In a typical DAY, how many hours do you spend watching TV? (Put 0 if you do not spend any time doing it)
    - Choices: Numpad, less than an hour a day, do not know, prefer not to answer
- Length of mobile phone use (fieldID 1110)
    - Question: For approximately how many years have you been using a mobile phone at least once per week to make or receive calls?
    - Choices: Never used mobile phone at least once per week, one year or less, two to four years, five to eight years, more than eight years, do not know, prefer not to answer
- Plays computer games (fieldID 2237)
    - Question: Do you play computer games?
    - Choices: Never/rarely, sometimes, often, prefer not to answer
- Sleep duration (fieldID 1160)
    - Question: About how many hours sleep do you get in every 24 hours? (please include naps)
    - Choices: Numpad, do not know, prefer not to answer
- Getting up in morning (fieldID 1170)
    - Question: On an average day, how easy do you find getting up in the morning?
    - Choices: Not at all easy, not very easy, fairly easy, very easy, do not know, prefer not to answer
- Morning/evening person (chronotype) (fieldID 1180)
    - Question: Do you consider yourself to be?
    - Choices: Definitely a ‘morning’ person, more of a ’morning’ than ‘evening’ person, more of an ’evening’ than a ‘morning’ person, definitely an ‘evening’ person, do not know, prefer not to answer
- Nap during day (fieldID 1190)
    - Question: Do you have a nap during the day?
    - Choices: Never/rarely, sometimes, usually, prefer not to answer
- Sleeplessness / insomnia (fieldID 1200)
    - Question: Do you have trouble falling asleep at night or do you wake up in the middle of the night?
    - Choices: Never/rarely, sometimes, usually, prefer not to answer
- Daytime dozing / sleeping (fieldID 1220)
    - Question: How likely are you to doze off or fall asleep during the daytime when you don't mean to? (e.g. when working, reading or driving)
    - Choices: Never/rarely, sometimes, often, all of the time, do not know, prefer not to answer
- Smoking status (fieldID 20116)
    - Derived from Current tobacco smoking (fieldID 1239) and Past tobacco smoking (fieldID 1249)
    - Classified as Never (0) if Current tobacco smoking = prefer not to say (-3) or no (0) AND Past tobacco smoking = just tried once or twice (3) or I have never smoked (4).

    - Classified as Previous (1) if Current tobacco smoking = no (0) AND Past tobacco smoking = smoked on most or all days (1) or smoked occasionally (2).
    - Classified as Current (2) if Current tobacco smoking = yes, on most or all days (1) or only occasionally (2).
    - Classified as Prefer not to say (-3) if either of the following conditions is met:
    - Current tobacco smoking = prefer not to say (-3) AND Past tobacco smoking = smoked on most or all days (1) or smoked occasionally (2).
    - Current tobacco smoking = no (0) AND Past tobacco smoking = prefer not to say (-3).
- Pack years of smoking (fieldID 20161)
    - Derived from other fields
    - Number of cigarettes per day / 20 * (Age stopped smoking - Age start smoking)
    - The figure is adjusted for individuals who gave up smoking for more than 6 months, according to data fields 2907 or 3486, by subtracting six months from the number of years of smoking (Number of cigarettes per day / 20 * (Age stopped smoking - Age start smoking - 0.5))
- Alcohol drinker status (fieldID 20117)
    - Derived from other fields
    - Choices: Never, previous, current, prefer not to answer
- Alcohol intake frequency (fieldID 1558)
    - Question: About how often do you drink alcohol?
    - Choices: Daily or almost daily, three or four times a week, once or twice a week, one to three times a month, special occasions only, never, prefer not to answer
- Time spent outdoors in summer (fieldID 1050)
    - Question: In a typical DAY in summer, how many hours do you spend outdoors?
    - Choices: Numpad, less than an hour a day, do not know, prefer not to answer
- Time spent outdoors in winter (fieldID 1060)
    - Question: In a typical DAY in winter, how many hours do you spend outdoors?
    - Choices: Numpad, less than an hour a day, do not know, prefer not to answer
- Use of sun/uv protection (fieldID 2267)
    - Question: Do you wear sun protection (e.g. sunscreen lotion, hat) when you spend time outdoors in the summer?
    - Choices: Never/rarely, sometimes, most of the time, always, do not go out in sunshine, do not know, prefer not to answer

**Social**

- Age first had sexual intercourse (fieldID 2139)
    - Question: What was your age when you first had sexual intercourse? (Sexual intercourse includes vaginal, oral or anal intercourse)
    - Choices: Numpad, never had sex, prefer not to answer, do not know

- Lifetime number of sexual partners (fieldID 2149)
    - Question: About how many sexual partners have you had in your lifetime?
    - Choices: Numpad, do not know, prefer not to answer
- Frequency of friend/family visits (fieldID 1031)
    - Question: How often do you visit friends or family or have them visit you?
    - Choices: Almost daily, 2-4 times a week, about once a week, about once a month, once every few months, never or almost never, no friends/family outside household, do not know, prefer not to answer
- Leisure/social activities (fieldID 6160)
    - Question: Which of the following do you attend once a week or more often? (You can select more than one)
    - Choices: Sports club or gym, pub or social club, religious group, adult education class, other group activity, none of the above, prefer not to answer
- Able to confide (fieldID 2110)
    - Question: How often are you able to confide in someone close to you?
    - Choices: Almost daily, 2-4 times a week, about once a week, about once a month, once every few months, never or almost never, do not know, prefer not to answer
- Family relationship satisfaction (fieldID 4559)
    - Question: In general how satisfied are you with your FAMILY RELATIONSHIPS?
    - Choices: Extremely happy, very happy, moderately happy, moderately unhappy, very unhappy, extremely unhappy, do not know, prefer not to answer
- Friendships satisfaction (fieldID 4570)
    - Question: In general how satisfied are you with your FRIENDSHIPS?
    - Choices: Extremely happy, very happy, moderately happy, moderately unhappy, very unhappy, extremely unhappy, do not know, prefer not to answer
- Number in household (fieldID 709)
    - Question: Including yourself, how many people are living together in your household? (Include those who usually live in the house such as students living away from home during term, partners in the armed forces or professions such as pilots)
    - Choices: Numpad, do not know, prefer not to answer

**Socioeconomic**

- Current employment status (fieldID 6142)
    - Question: Which of the following describes your current situation? (You can select more than one answer)
    - Choices: In paid employment or self-employed, retired, looking after home and/or family, unable to work because of sickness or disability, unemployed, doing unpaid or voluntary work, full or part-time student, none of the above, prefer not to answer
- Length of working week for main job (fieldID 767)

- Question: In a typical WEEK, how many hours do you spend at work? (Do not include hours travelling to and from work)
    - Choices: Numpad, do not know, prefer not to answer
- Job involves mainly walking or standing (fieldID 806)
    - Question: Does your work involve walking or standing for most of the time?
    - Choices: Never/rarely, sometimes, usually, always, do not know, prefer not to answer
- Job involves heavy manual or physical work (fieldID 816)
    - Question: Does your work involve heavy manual or physical work?
    - Choices: Never/rarely, sometimes, usually, always, do not know, prefer not to answer
- Job involves shift work (fieldID 826)
    - Question: Does your work involve shift work?
    - Choices: Never/rarely, sometimes, usually, always, do not know, prefer not to answer
- Qualifications (fieldID 6138)
    - Question: Which of the following qualifications do you have? (You can select more than one)
    - Choices: College or University degree, A levels/AS levels or equivalent, O levels/GCSEs or equivalent, CSEs or equivalent, NVQ or HND or HNC or equivalent, other professional qualifications eg: nursing, teaching, none of the above, prefer not to answer
- Private healthcare (fieldID 4674)
    - Question: Do you use private healthcare?
    - Choices: Yes all of the time, yes most of the time, yes sometimes, no never, do not know, prefer not to answer
- Financial situation satisfaction (fieldID 4581)
    - Question: In general how satisfied are you with your FINANCIAL SITUATION?
    - Choices: Extremely happy, very happy, moderately happy, moderately unhappy, very unhappy, extremely unhappy, do not know, prefer not to answer
- Work/job satisfaction (fieldID 4537)
    - Question: In general how satisfied are you with the WORK that you do?
    - Choices: Extremely happy, very happy, moderately happy, moderately unhappy, very unhappy, extremely unhappy, I am not employed, do not know, prefer not to answer
- Health satisfaction (fieldID 4548)
    - Question: In general how satisfied are you with your HEALTH?
    - Choices: Extremely happy, very happy, moderately happy, moderately unhappy, very unhappy, extremely unhappy, do not know, prefer not to answer

#### Supplementary Methods 2. Brain MRI processing for deformation-based morphometry

To obtain voxel-wise measures of brain morphometry via deformation-based morphometry, we processed the brain MRI data using a custom pipeline developed by our group to maximize registration accuracy.[1] T1w and FLAIR images were denoised,[2] corrected for N3 inhomogeneities,[3] and intensity-normalized. Brain masks were calculated using the BEaST algorithm.[4] A UKBB study-specific template space was generated using the processed T1w images of 200 subjects with representative age and sex distributions (available at https://github.com/CoBrALab/WMH_patho_UKB). All modalities were aligned at the subject level by performing rigid registration of other modalities (FLAIR and diffusion) to T1w subject space using the Advanced Normalization Tools software.[5] We then performed multispectral non-linear registration to the study-specific templates using preprocessed T1w images and fractional anisotropy maps supersampled to 1 mm isotropic[6] as inputs.[5] This multispectral registration scheme enhances registration accuracy, especially in white matter regions.[7]

### Supplementary Tables

| Category | Field ID | Name | Transformation | Logical imputation (before imputing across timepoints) | Missing values (TP0) | Missing values (TP1) | Missing values (TP2) | Missing values (TP3) |
|---|---|---|---|---|---|---|---|---|
| Diet | 1289 | Cooked Vegetables | Less than one = 0 | | 767 (0.2%) | 0 (0%) | 0 (0%) | 0 (0%) |
| Diet | 1299 | Raw Vegetables | Less than one = 0 | | 755 (0.2%) | 12 (0.1%) | 18 (0%) | 0 (0%) |
| Diet | 1309 | Fresh Fruits | Less than one = 0 | | 146 (0%) | 418 (2.1%) | 899 (1.5%) | 41 (0.8%) |
| Diet | 1319 | Dried Fruits | Less than one = 0 | | 912 (0.3%) | 0 (0%) | 0 (0%) | 0 (0%) |
| Diet | 1329 | Oily Fish | Made continuous (0=Never, 5=Once or more daily) | | 398 (0.1%) | 0 (0%) | 2 (0%) | 0 (0%) |
| Diet | 1339 | Non-oily Fish | Made continuous (0=Never, 5=Once or more daily) | | 329 (0.1%) | 2 (0%) | 0 (0%) | 0 (0%) |
| Diet | 1349 | Processed Meat | Made continuous (0=Never, 5=Once or more daily) | | 86 (0%) | 16 (0.1%) | 23 (0%) | 0 (0%) |
| Diet | 1359 | Poultry | Made continuous (0=Never, 5=Once or more daily) | | 60 (0%) | 734 (3.6%) | 1653 (2.7%) | 59 (1.1%) |
| Diet | 1369 | Beef | Made continuous (0=Never, 5=Once or more daily) | | 252 (0.1%) | 76 (0.4%) | 155 (0.3%) | 1 (0%) |
| Diet | 1379 | Lamb/Mutton | Made continuous (0=Never, 5=Once or more daily) | | 374 (0.1%) | 1463 (7.2%) | 3727 (6.1%) | 171 (3.2%) |
| Diet | 1389 | Pork | Made continuous (0=Never, 5=Once or more daily) | | 363 (0.1%) | 54 (0.3%) | 141 (0.2%) | 3 (0.1%) |
| Diet | 1408 | Cheese | Made continuous (0=Never, 5=Once or more daily) | | 4347 (1.4%) | 4750 (23.4%) | 12766 (20.9%) | 742 (14%) |
| Diet | 1438 | Bread | Less than one = 0 | | 639 (0.2%) | 2 (0%) | 14 (0%) | 1 (0%) |
| Diet | 1458 | Cereal | Less than one = 0 | | 165 (0.1%) | 188 (0.9%) | 405 (0.7%) | 12 (0.2%) |
| Diet | 1478 | Salt added | Made continuous (0=Never/rarely, 3=Always) | | 5 (0%) | 156 (0.8%) | 365 (0.6%) | 13 (0.2%) |
| Diet | 1488 | Tea | Less than one = 0 | | 58 (0%) | 2 (0%) | 7 (0%) | 1 (0%) |
| Diet | 1498 | Coffee | Less than one = 0 | | 74 (0%) | 5 (0%) | 11 (0%) | 1 (0%) |
| Diet | 1528 | Water | Less than one = 0 | | 474 (0.2%) | 21 (0.1%) | 50 (0.1%) | 0 (0%) |
| Diet | 1548 | Variations in Diet | Made continuous (0=Never/rarely, 2=Sometimes) | | 234 (0.1%) | 17 (0.1%) | 22 (0%) | 0 (0%) |
| Lifestyle | 864 | Days/Week Walking | Unable to walk = 0 | | 857 (0.3%) | 6 (0%) | 13 (0%) | 2 (0%) |
| Lifestyle | 874 | Duration Walks | | | 18748 (6.1%) | 0 (0%) | 2 (0%) | 1 (0%) |
| Lifestyle | 884 | Days/Week Moderate Activity | | | 4496 (1.5%) | 843 (4.2%) | 2234 (3.7%) | 133 (2.5%) |
| Lifestyle | 894 | Duration Moderate Activity | | | 36160 (11.7%) | 1 (0%) | 1 (0%) | 0 (0%) |
| Lifestyle | 904 | Days/Week Vigorous Activity | | | 4660 (1.5%) | 1 (0%) | 1 (0%) | 0 (0%) |
| Lifestyle | 914 | Duration Vigorous Activity | | | 87388 (28.3%) | 1 (0%) | 4 (0%) | 0 (0%) |
| Lifestyle | 1050 | Time Outdoors (Summer) | Less than an hour a day = 0 | | 5195 (1.7%) | 4 (0%) | 27 (0%) | 1 (0%) |
| Lifestyle | 1060 | Time Outdoors (Winter) | Less than an hour a day = 0 | | 5103 (1.7%) | 5 (0%) | 18 (0%) | 1 (0%) |
| Lifestyle | 1070 | Time Watching TV | Less than an hour a day = 0 | | 343 (0.1%) | 2 (0%) | 2 (0%) | 0 (0%) |
| Lifestyle | 1080 | Time on Computer | Less than an hour a day = 0 | | 594 (0.2%) | 10 (0%) | 12 (0%) | 0 (0%) |
| Lifestyle | 1090 | Time Driving | Less than an hour a day = 0 | | 1515 (0.5%) | 2 (0%) | 5 (0%) | 0 (0%) |
| Lifestyle | 1110 | Length Mobile Phone Use | Made continuous (0=Never used mobile phone at least once per week, 4=More than eight years) | | 1372 (0.4%) | 1 (0%) | 3 (0%) | 0 (0%) |
| Lifestyle | 1160 | Sleep Duration | | | 333 (0.1%) | 1 (0%) | 1 (0%) | 0 (0%) |
| Lifestyle | 1170 | Ease Getting up in Morning | Made continuous (0=Not at all easy, 3=Very easy) | | 93 (0%) | 1 (0%) | 0 (0%) | 0 (0%) |
| Lifestyle | 1180 | Morning Person | Made continuous (0=Definitely an 'evening' person, 3=Definitely a 'morning' person) | | 17809 (5.8%) | 2 (0%) | 6 (0%) | 0 (0%) |
| Lifestyle | 1190 | Nap During Day | Made continuous (0=Never/rarely, 2=Usually) | | 78 (0%) | 3 (0%) | 6 (0%) | 0 (0%) |
| Lifestyle | 1200 | Sleeplessness | Made continuous (0=Never/rarely, 2=Usually) | | 67 (0%) | 1 (0%) | 11 (0%) | 0 (0%) |
| Lifestyle | 1220 | Daytime Sleepiness | Made continuous (0=Never/rarely, 3=All of the time) | | 359 (0.1%) | 158 (0.8%) | 367 (0.6%) | 27 (0.5%) |
| Lifestyle | 1558 | Alcohol Intake Freq. | Made continuous (0=Never, 5=Daily or almost daily) | | 37 (0%) | 16 (0.1%) | 35 (0.1%) | 3 (0.1%) |
| Lifestyle | 2237 | Play Computer Games | Made continuous (0=Never/rarely, 2=Often) | | 112 (0%) | 0 (0%) | 2 (0%) | 0 (0%) |
| Lifestyle | 2267 | Use UV Protection | Made continuous (0=Do not go out in sunshine, 4=Always) | | 163 (0.1%) | 0 (0%) | 0 (0%) | 0 (0%) |
| Lifestyle | 20116 | Smoking Status | Made continuous (0=Never, 1=Previous, 2=Current) | | 337 (0.1%) | 0 (0%) | 1 (0%) | 0 (0%) |
| Lifestyle | 20117 | Alcohol Drinker Status | Made continuous (0=Never, 1=Previous, 2=Current) | | 64 (0%) | 0 (0%) | 0 (0%) | 0 (0%) |
| Lifestyle | 20161 | Pack Years Smoking | | if is(NA) & smoking status == "Never", 0 | 32359 (10.5%) | 6 (0%) | 21 (0%) | 0 (0%) |

| | | | | | | | | |
|---|---|---|---|---|---|---|---|---|
| Social | 709 | Number People in Household | | | 780 (0.3%) | 2 (0%) | 4 (0%) | 0 (0%) |
| Social | 1031 | Freq. Friend/Family Visits | Made continuous (0=Never or almost never OR No friends/family outside household, 5=Almost daily) | | 395 (0.1%) | 0 (0%) | 0 (0%) | 0 (0%) |
| Social | 2110 | Able to Confide | Made continuous (0=Never or almost never, 5=Almost daily) | | 4699 (1.5%) | 94 (0.5%) | 252 (0.4%) | 7 (0.1%) |
| Social | 2139 | Age at First Intecourse | Never had sex = 99 | | 18898 (6.1%) | 903 (4.5%) | 2370 (3.9%) | 148 (2.8%) |
| Social | 2149 | Lifetime Sexual Partners | | | 32895 (10.7%) | 1681 (8.3%) | 4927 (8.1%) | 335 (6.3%) |
| Social | 4559 | Family Relationship Satisfaction | Made continuous (0=Extremely unhappy, 5=Extremely happy) | | 97745 (31.7%) | 1 (0%) | 0 (0%) | 0 (0%) |
| Social | 4570 | Friendship Satisfaction | Made continuous (0=Extremely unhappy, 5=Extremely happy) | | 98097 (31.8%) | 0 (0%) | 3 (0%) | 0 (0%) |
| Social | 6160 | Leisure/social activities | Each choice dichotomized into Yes/No | | 0 (0%) | 8657 (42.7%) | 27462 (45%) | 2319 (43.7%) |
| Socioeconomic | 680 | Own accomodation | Dichotomized (1=Own outright OR Own with a martgage) | | 0 (0%) | 27 (0.1%) | 210 (0.3%) | 3 (0.1%) |
| Socioeconomic | 767 | Length of Work Week | | if is(NA) & employement status == "Retired", 0 | 19472 (6.3%) | 104 (0.5%) | 452 (0.7%) | 15 (0.3%) |
| Socioeconomic | 806 | Standing/walking Job | Made continuous (0=Never/rarely, 3=Always) | If is(NA) & employement status != "In paid employment or self-employed", 0 | 52 (0%) | 131 (0.6%) | 587 (1%) | 17 (0.3%) |
| Socioeconomic | 816 | Physical Job | Made continuous (0=Never/rarely, 3=Always) | If is(NA) & employement status != "In paid employment or self-employed", 0 | 50 (0%) | 37 (0.2%) | 212 (0.3%) | 1 (0%) |
| Socioeconomic | 826 | Shift Job | Made continuous (0=Never/rarely, 3=Always) | If is(NA) & employement status != "In paid employment or self-employed", 0 | 135 (0%) | 52 (0.3%) | 270 (0.4%) | 4 (0.1%) |
| Socioeconomic | 4537 | Job Satisfaction | Made continuous (0=Extremely unhappy, 6=Extremely happy, 3=Unemployed) | | 171059 (55.4%) | 0 (0%) | 0 (0%) | 0 (0%) |
| Socioeconomic | 4548 | Health Satisfaction | Made continuous (0=Extremely unhappy, 5=Extremely happy) | | 96729 (31.4%) | 0 (0%) | 0 (0%) | 0 (0%) |
| Socioeconomic | 4581 | Finances Satisfaction | Made continuous (0=Extremely unhappy, 5=Extremely happy) | | 96845 (31.4%) | 0 (0%) | 0 (0%) | 0 (0%) |
| Socioeconomic | 4674 | Access Private Healthcare | Made continuous (0=No/never, 3=Yes/all of the time) | | 97198 (31.5%) | 4 (0%) | 15 (0%) | 1 (0%) |
| Socioeconomic | 6138 | Qualifications | Each choice dichotomized into Yes/No | | 0 (0%) | 0 (0%) | 0 (0%) | 0 (0%) |
| Socioeconomic | 6142 | Current employment status | Each choice dichotomized into Yes/No | | 0 (0%) | 1840 (9.1%) | 5650 (9.3%) | 367 (6.9%) |

#### Supplementary Table 1. Input variable transformations, imputations, and missing values

All variables used as inputs to the CatBoost models are shown with their field IDs, transformations, logical imputation rules, and missing values across timepoints. Logical imputation rules were applied before imputation across timepoints. Missing values were calculated in the final sample after excluding participants with >10% missing values and are equivalent to those that were later computationally imputed.

| Data type | UKB categories | TP | Sample size range | Model |
|---|---|---|---|---|
| Blood biomarkers | 17518, 100081 | 0 | 24,913-290,543 | Phenotype ~ IGL + bs(age) + height + weight + bs(day of year) + bs(time of day) + (1\|assessment center) |
| Abdominal MRI | 158, 156, 126 | 2 | 27,761-39,229 | Phenotype ~ IGL + bs(age) + height + weight + (1\|assessment center) |
| DXA scan | 124 | 2 | 15,831-45,065 | Phenotype ~ IGL + bs(age) + height + weight + (1\|assessment center) |
| Arterial stiffness | 100007 | 0 | 155,462-155,463 | Phenotype ~ IGL + bs(age) + (1\|assessment center) + height + (1\|device ID) |
| Blood pressure | 100011 | 0 | 280,284-280,293 | Phenotype ~ IGL + bs(age) + (1\|assessment center) + height + pulse rate + (1\|device ID) |
| ECG exercise<br>ECG rest | 100012<br>104 | 0<br>2 | 63,183-63,195<br>41,040-50,406 | Phenotype ~ IGL + bs(age) + (1\|assessment center) + height<br>Phenotype ~ IGL + bs(age) + (1\|assessment center) + height + weight + (1\|device ID) |
| Heel bone density | 100018 | 0 | 121,785-121,832 | Phenotype ~ IGL + bs(age) + (1\|assessment center) + height + weight + (1\|device ID) |
| Hand grip strength | 100019 | 0 | 298,332 | Phenotype ~ IGL + bs(age) + (1\|assessment center) + height + (1\|device ID) |
| Spirometry | 100020 | 0 | 170,057 | Phenotype ~ IGL + bs(age) + (1\|assessment center) + height + (1\|device ID) + smoked cigarette within last hour + drank caffeine within last hour + used an inhaler within last hour |
| Hearing test | 100049 | 0 | 147,951-147,959 | Phenotype ~ IGL + bs(age) + (1\|assessment center) + duration of hearing test + volume level set by participant |
| Carotid ultrasound | 101 | 2 | 46,911-47,568 | Phenotype ~ IGL + bs(age) + (1\|assessment center) + height + weight |
| Brain imaging<br>Brain imaging (voxel) | 508<br>- | 2<br>2 | 32,776-35,024<br>33,008 | Phenotype ~ IGL + bs(Age) + intracranial volume + motion t1 + scanner X + scanner Y + scanner Z + scanner position + (1\|assessment center)<br>Phenotype ~ IGL + bs(Age) + motion t1 + scanner X + scanner Y + scanner Z + scanner position + (1\|assessment center) |
| Cognition | 100026 | 2 | 38,712-54,313 | Phenotype ~ IGL + bs(age) + (1\|assessment center) |
| Mental health | 100060 | 0 | 156,693-296,613 | Phenotype ~ IGL + bs(age) + (1\|assessment center) |
| Medical diagnoses | 1712 | 0 | 301,113 | Group (cases vs controls) ~ IGL + age |
| Medication | 100075 | 0 | 301,113 | Medication use (cases vs controls) ~ IGL + age |

#### Supplementary Table 2. Models used for associations between the IGL and health phenotypes

For each type of health phenotype, the statistical model used with all covariates is shown. Timepoint (TP), Index of Gendered Life-domains (IGL), B-spline (bs), magnetic resonance imaging (MRI), dual-energy X-ray absorptiometry (DXA), electrocardiogram (ECG).

### Supplementary Figures

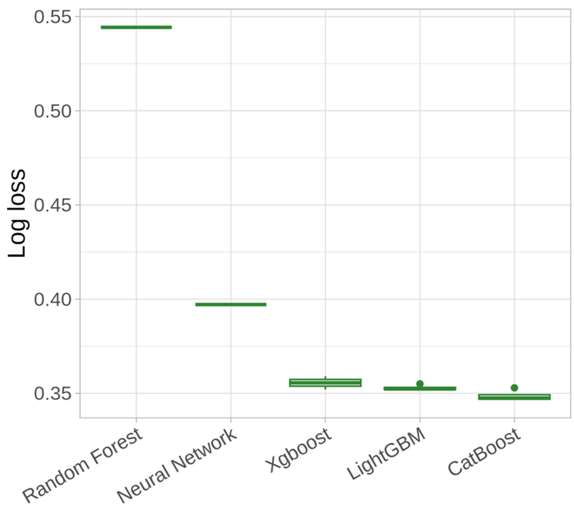

#### Supplementary Figure 1. AutoML performance across model types.

Using timepoint 0 data, different machine learning architectures were tested using the MLjar-Supervised AutoML Python package. The model with the lowest log loss value was a CatBoost model.

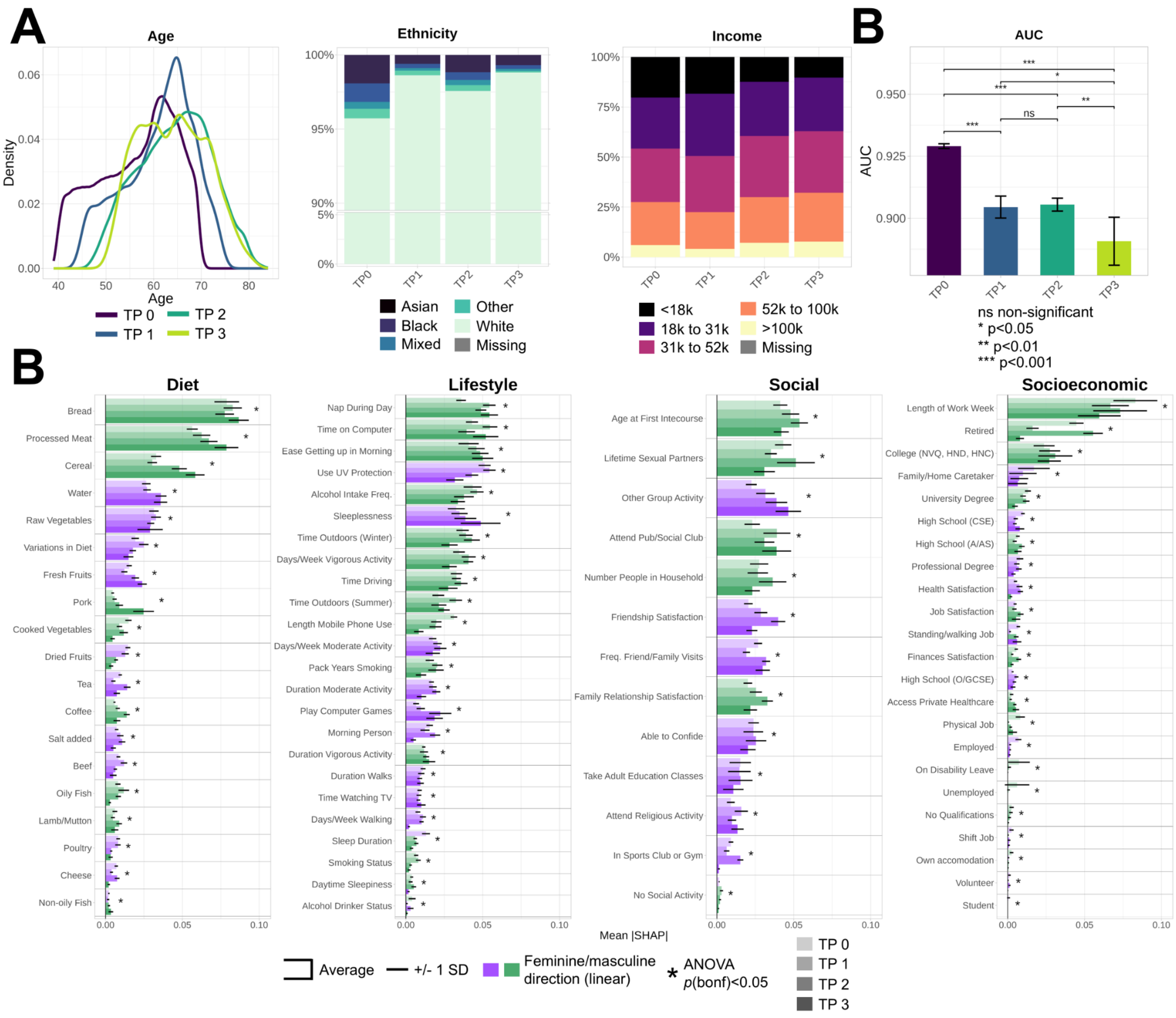

#### Supplementary Figure 2. Gendered dynamics across timepoints using complete, unmatched samples.

IGL models were computed at the four timepoints (TP) of the UKBB using the complete, unmatched samples. **A)** Distributions of age, ethnicity, and household income across timepoints. **B)** AUC values were compared with unpaired DeLong tests. **C)** Mean absolute SHAP variable importances across timepoints (from light to dark). Error bars indicate +/- one standard deviation (SD). A linear direction was calculated with a Pearson correlation on the averaged SHAP plot and is indicated by a light purple (feminine) or light green (masculine) color. Statistical differences between timepoints for each variable were calculated using ANOVAs, *p*-values were adjusted with a Bonferroni correction, and significant effects at p<0.05 are indicated with an asterisk.

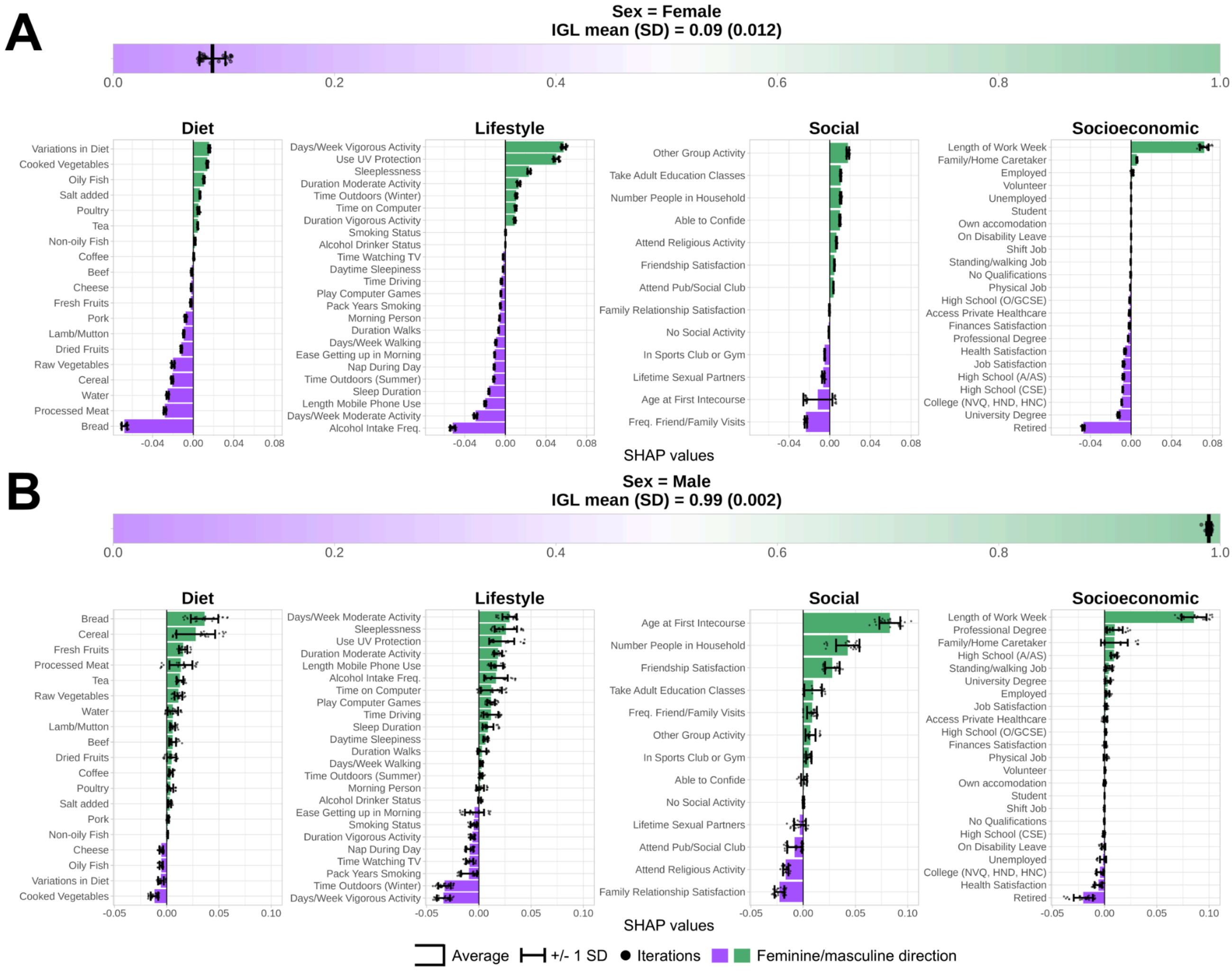

**Supplementary Figure 3. Examples of single-subject SHAP explanations for IGL-congruent individuals.**

Subject-specific SHAP values are shown for an IGL-congruent female **(A)** and male **(B)**. Top: the sex, IGL mean, and IGL standard deviation (SD) calculated across the 25 out-of-fold predictions are shown. Bottom: SHAP values explaining why each subject was assigned a specific IGL. Positive values indicate that the value of that variable shifted the prediction towards male (light green), and vice versa for female (light purple). The bars indicate the averaged SHAP value, while the error bars indicate +/- one SD.

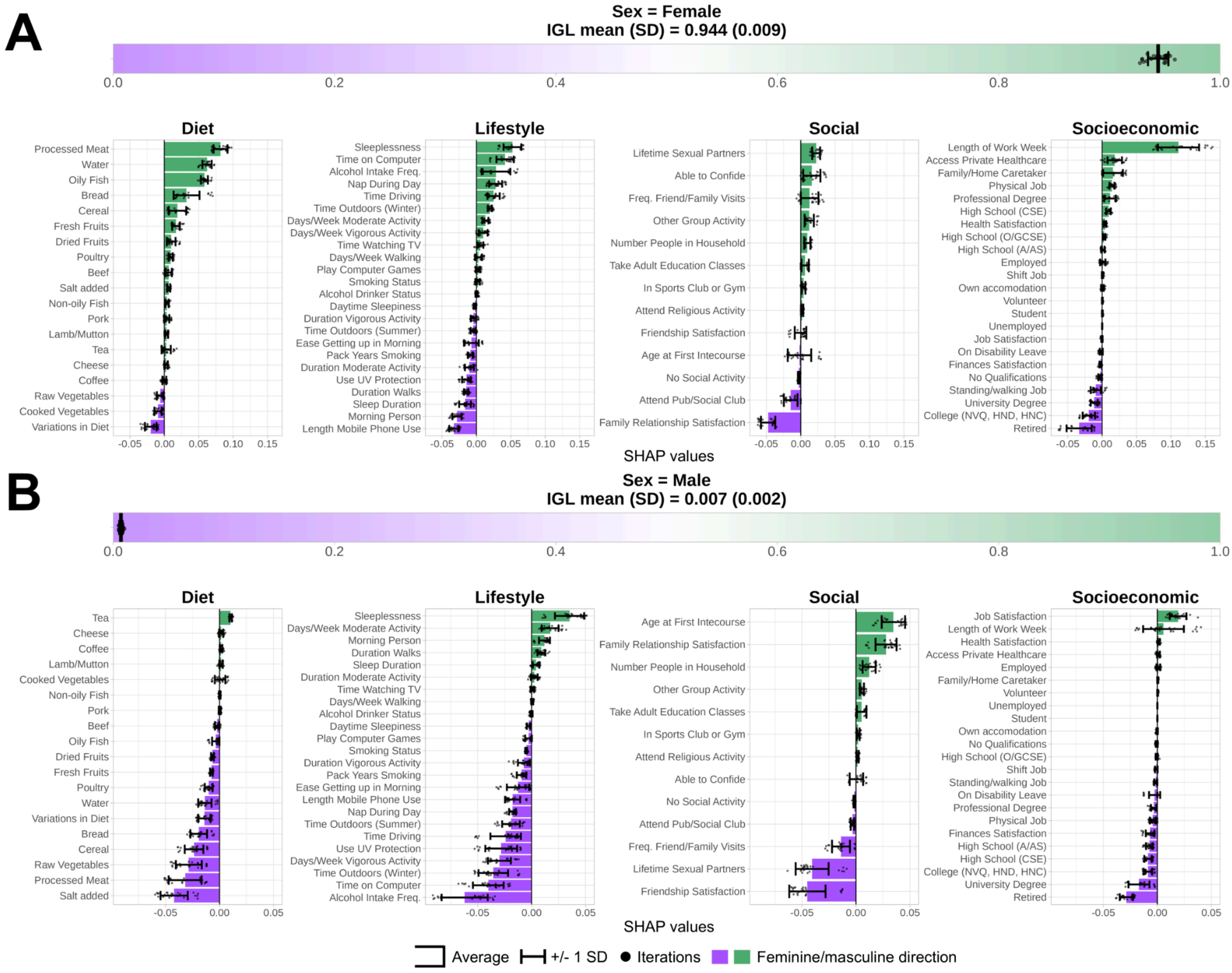

**Supplementary Figure 4. Examples of single-subject SHAP explanations for IGL-incongruent individuals.**

Subject-specific SHAP values are shown for an IGL-incongruent female **(A)** and male **(B)**. Top: the sex, IGL mean, and IGL standard deviation (SD) calculated across the 25 out-of-fold predictions are shown. Bottom: SHAP values explaining why each subject was assigned a specific IGL. Positive values indicate that the value of that variable shifted the prediction towards male (light green), and vice versa for female (light purple). The bars indicate the averaged SHAP value, while the error bars indicate +/- one SD.

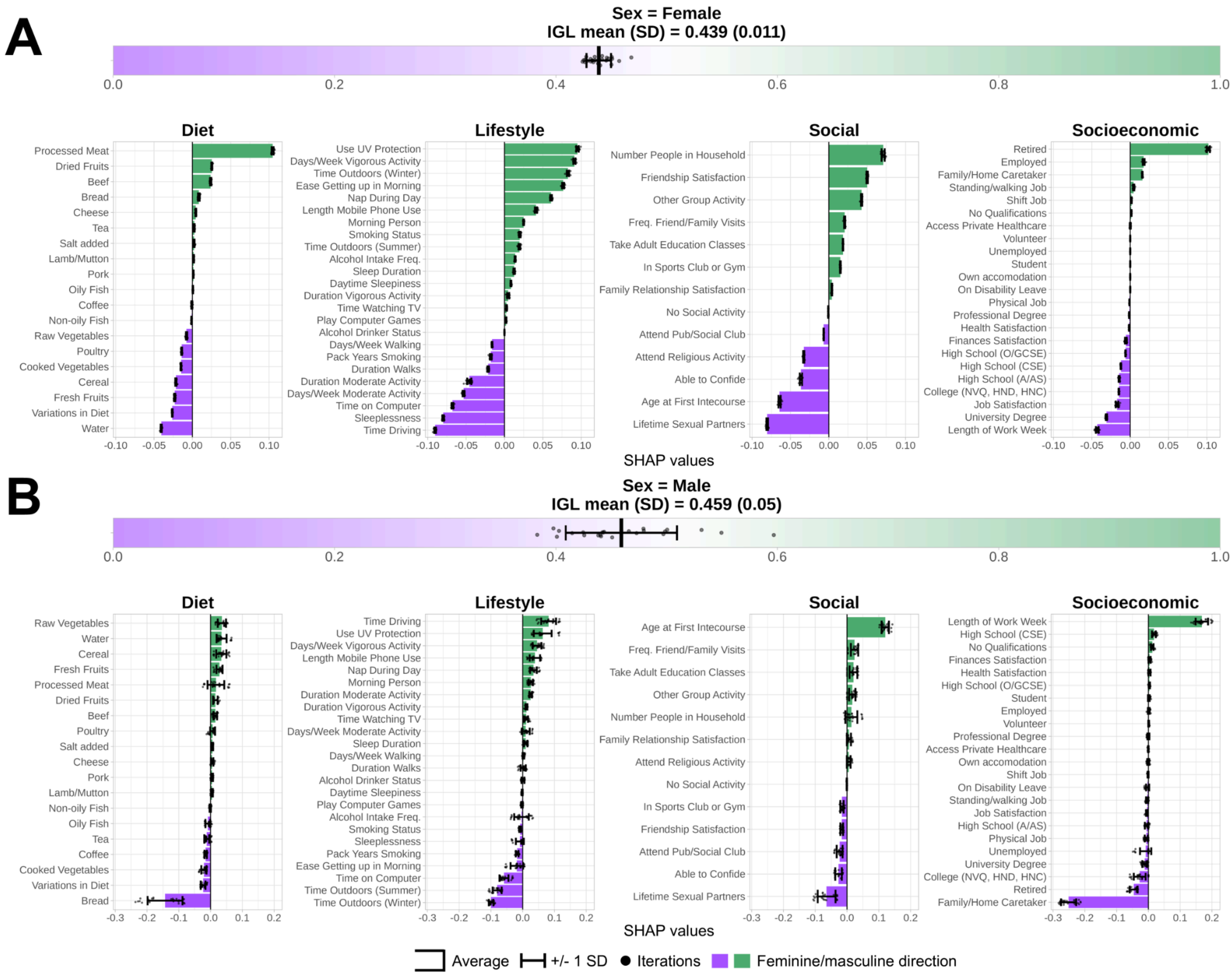

#### Supplementary Figure 5. Examples of single-subject SHAP explanations for IGL-androgynous individuals.

Subject-specific SHAP values are shown for an IGL-androgynous female **(A)** and male **(B)**. Top: the sex, IGL mean, and IGL standard deviation (SD) calculated across the 25 out-of-fold predictions are shown. Bottom: SHAP values explaining why each subject was assigned a specific IGL. Positive values indicate that the value of that variable shifted the prediction towards male (light green), and vice versa for female (light purple). The bars indicate the averaged SHAP value, while the error bars indicate +/- one SD.

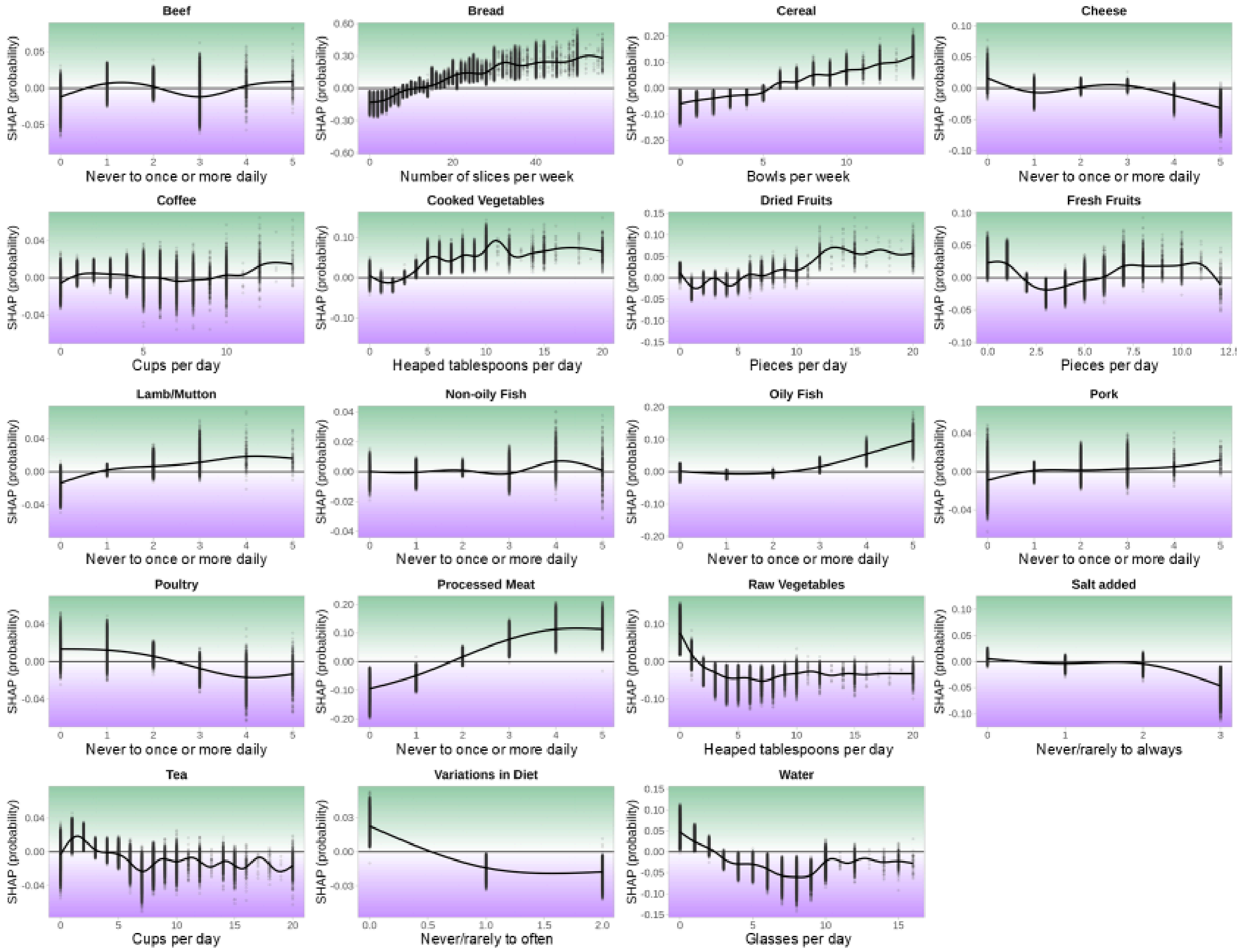

#### Supplementary Figure 6. Variable-level SHAP plots for diet predictors.

For continuous variables, large positive outliers above the 99.9th percentile were excluded for visualization purposes only. Generalized additive models were fit to the data using a varying number of basis dimensions (equivalent to the number of unique values up to 15).

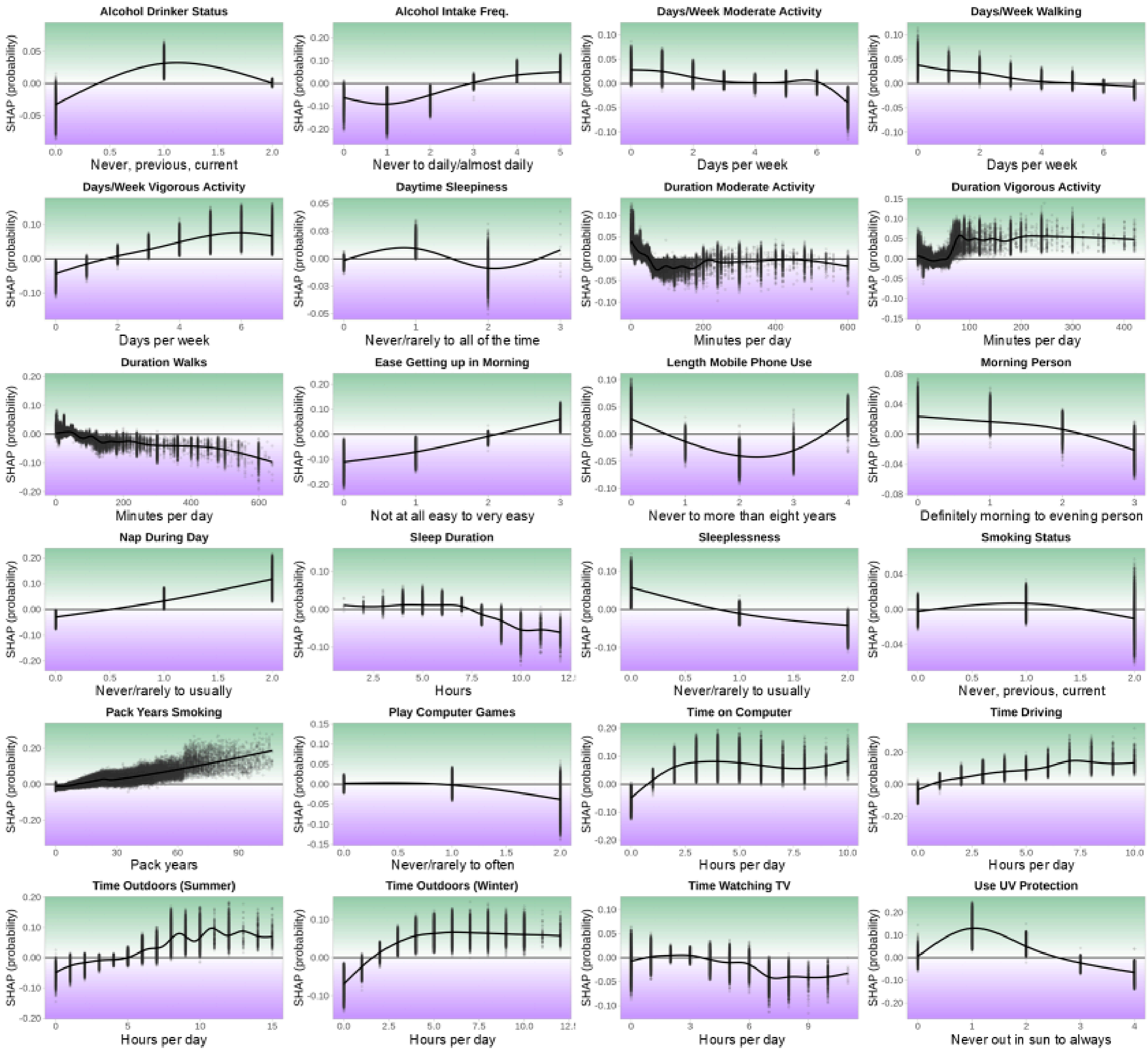

**Supplementary Figure 7. Variable-level SHAP plots for lifestyle predictors.**

For continuous variables, large positive outliers above the 99.9th percentile were excluded for visualization purposes only. Generalized additive models were fit to the data using a varying number of basis dimensions (equivalent to the number of unique values up to 15).

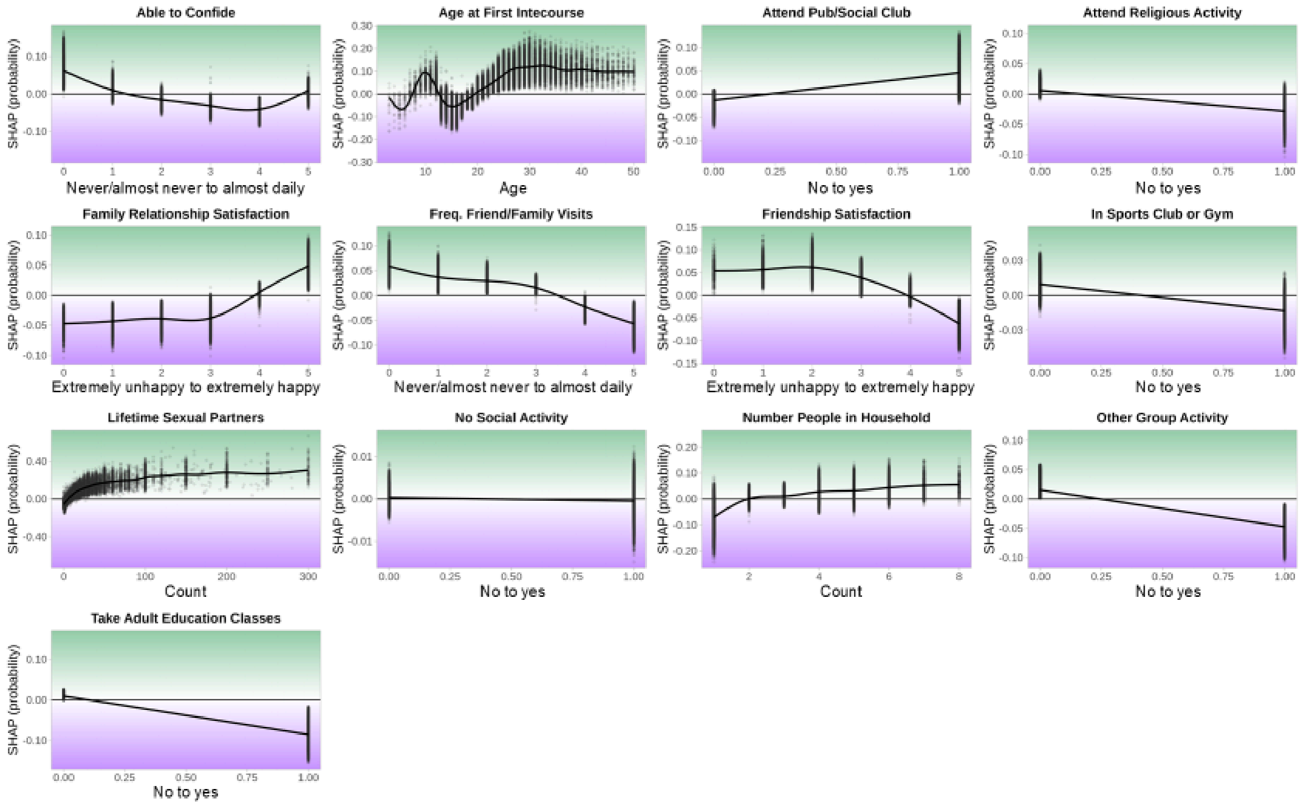

**Supplementary Figure 8. Variable-level SHAP plots for social predictors.**

For continuous variables, large positive outliers above the 99.9th percentile were excluded for visualization purposes only. Generalized additive models were fit to the data using a varying number of basis dimensions (equivalent to the number of unique values up to 15).

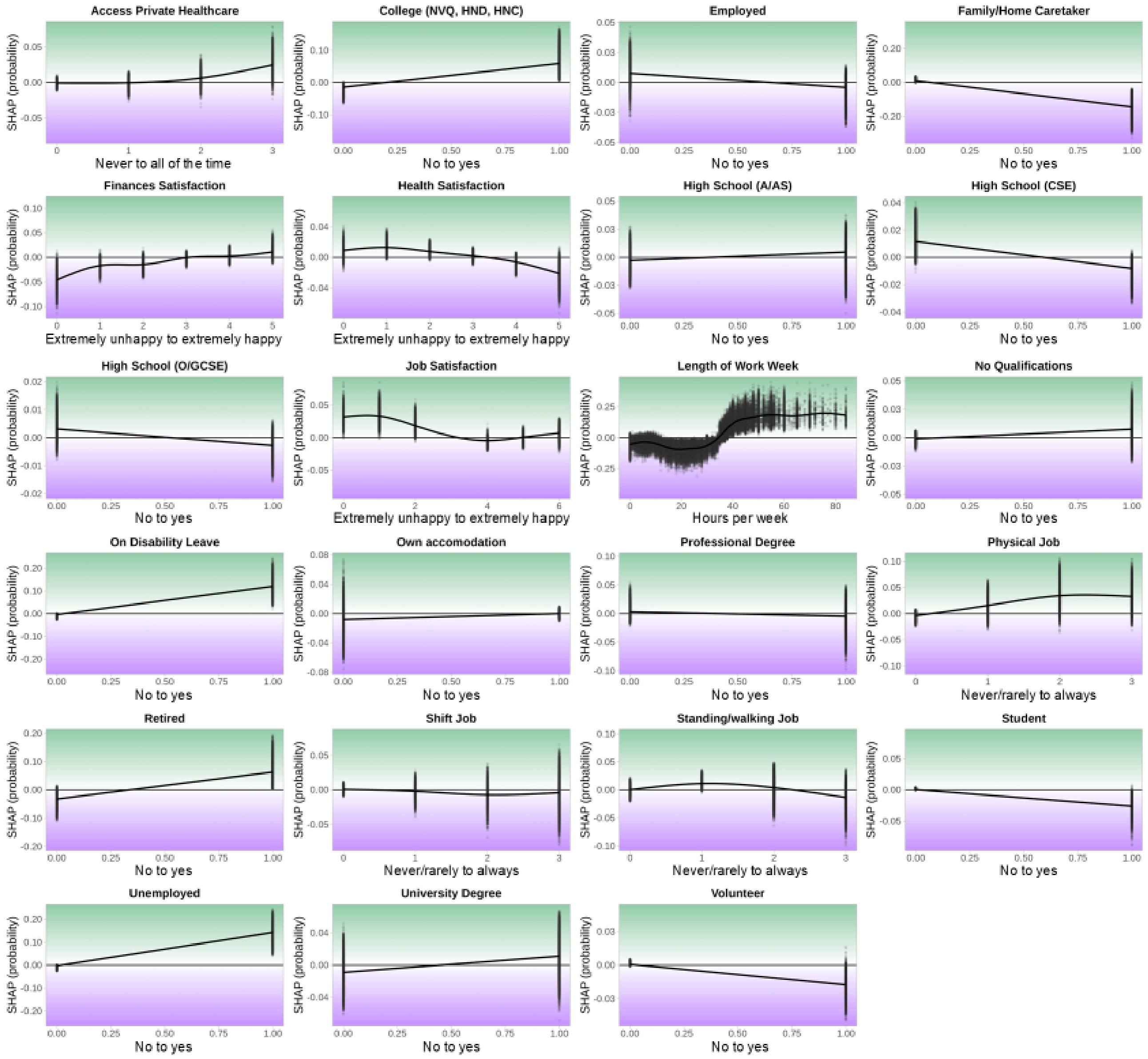

#### Supplementary Figure 9. Variable-level SHAP plots for socioeconomic predictors.

For continuous variables, large positive outliers above the 99.9th percentile were excluded for visualization purposes only. Generalized additive models were fit to the data using a varying number of basis dimensions (equivalent to the number of unique values up to 15).

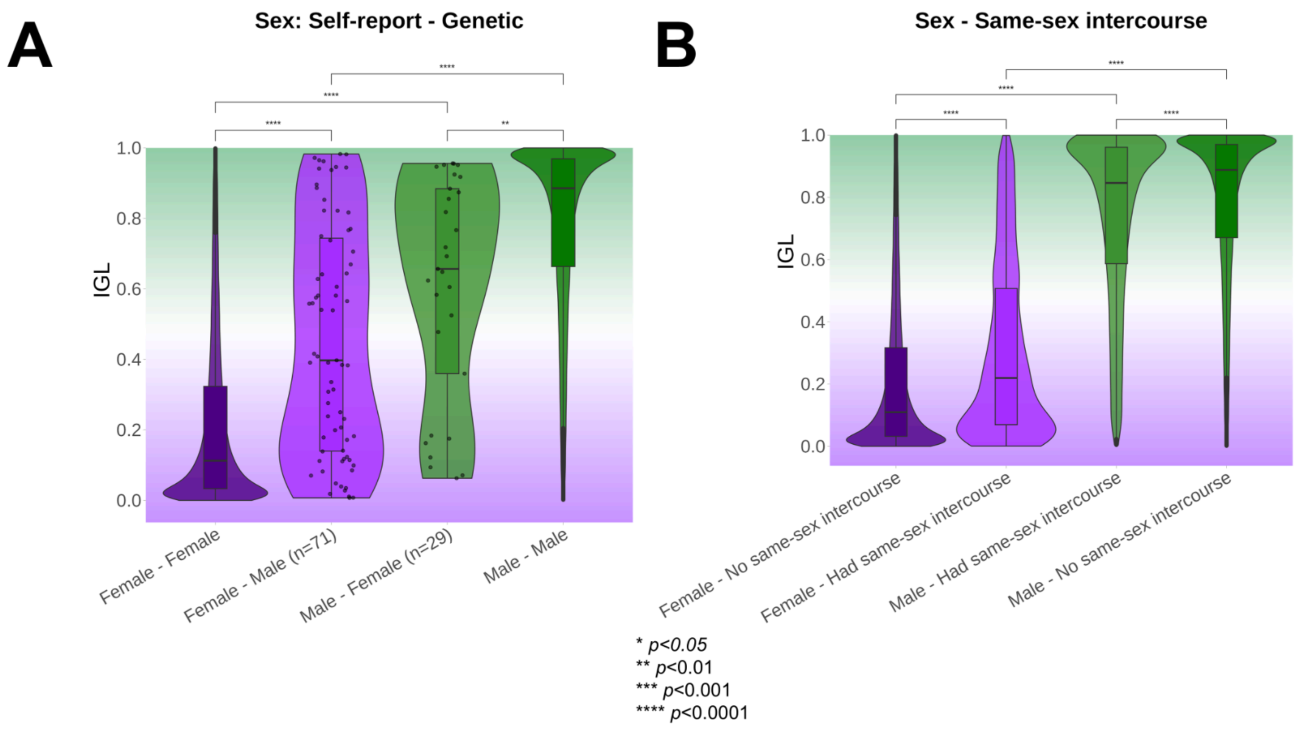

**Supplementary Figure 10. IGL distributions of sex-incongruent individuals and individuals who have had same-sex intercourse.**

Significant differences between distributions are indicated at different thresholds with asterisks. **A)** Individuals with an incongruent chromosomal and self-reported sex were held out of the training set and later predicted to estimate their IGL scores. Shown here are the distributions of sex-congruent and sex-incongruent participants. Both self-reported females with male chromosomes (*n*=71) and self-reported males with female chromosomes (*n*=29) without sex chromosome aneuploidies had more androgynous IGLs than sex-congruent individuals. **B)** Both females and males who reported ever having had same-sex intercourse had more androgynous IGLs than participants who did not.

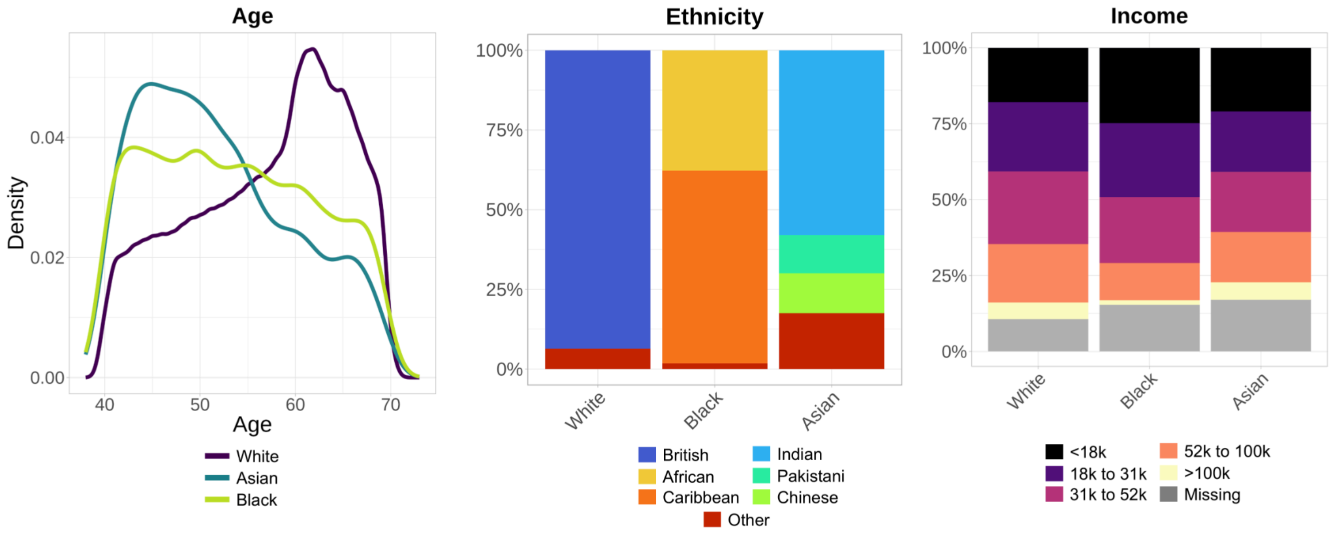

**Supplementary Figure 11. Demographics of ethnic groups before matching.**
Distributions of age, ethnicity subgroup, and household income across ethnicity groups before the matching procedure.

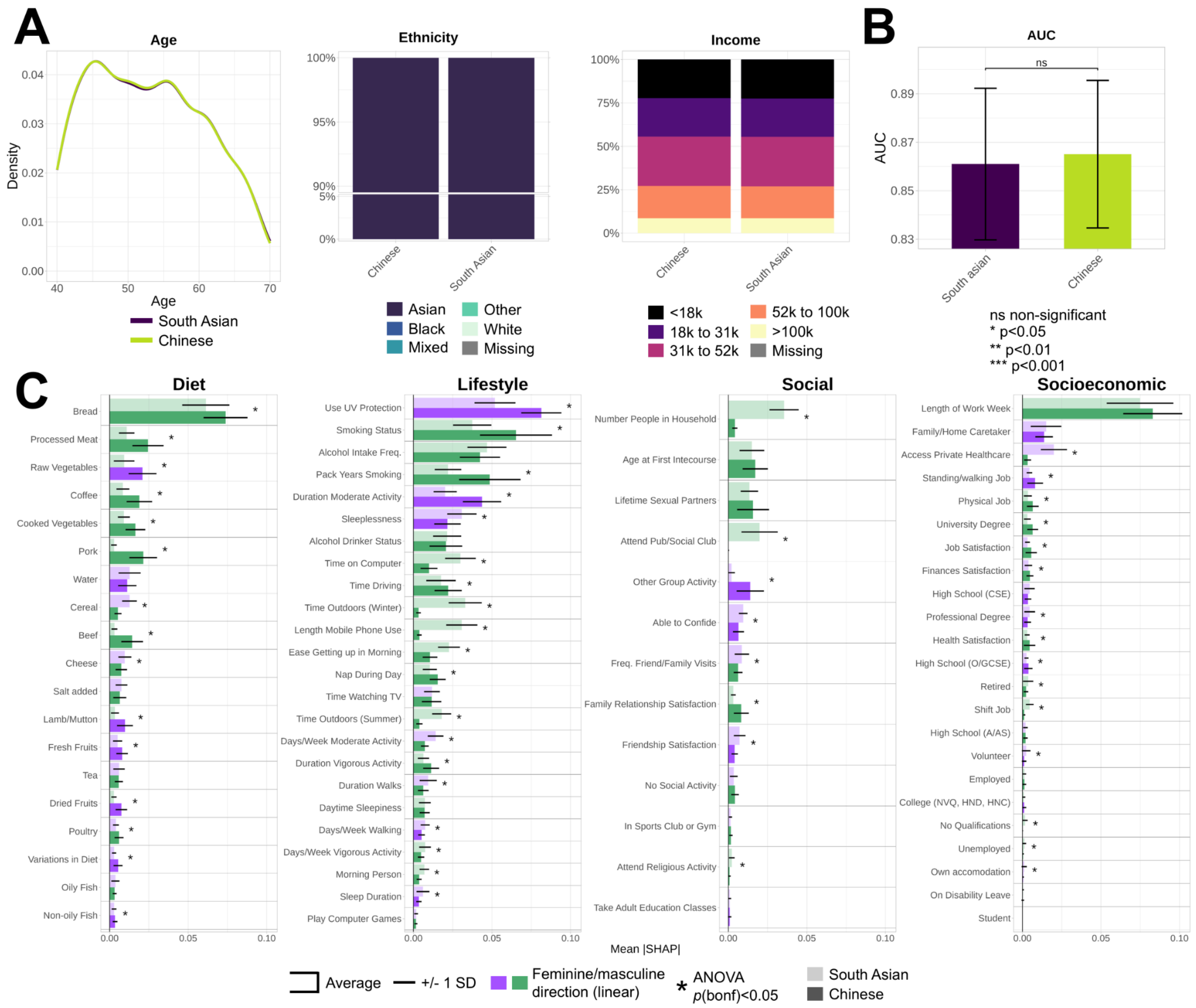

**Supplementary Figure 12. Gendered dynamics in South Asian and Chinese ethnicities.**

IGL models were computed separately in South Asian and Chinese ethnicities matched on age and household income. **A)** Distributions of age, ethnicity, and household income across Asian ethnicities. **B)** AUC values were compared with unpaired DeLong tests. **C)** Mean absolute SHAP variable importances across Asian ethnicities. Error bars indicate +/- one standard deviation (SD). A linear direction was calculated with a Pearson correlation on the averaged SHAP plot and is indicated by a light purple (feminine) or light green (masculine) color. Statistical differences between groups for each variable were calculated using ANOVAs, *p*-values were adjusted with a Bonferroni correction, and significant effects at $p<0.05$ are indicated with an asterisk.

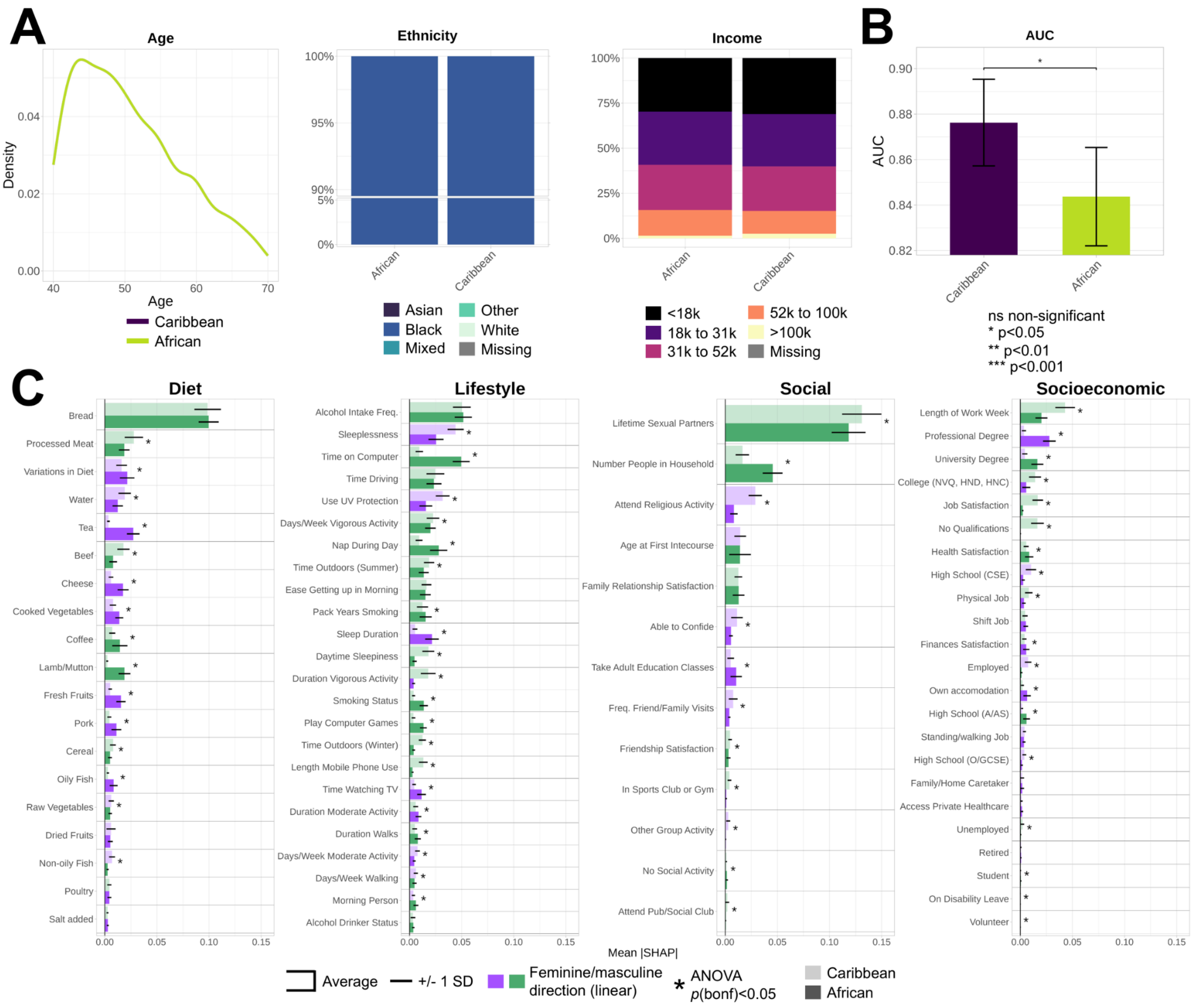

**Supplementary Figure 13. Gendered dynamics in Caribbean and African ethnicities.**

IGL models were computed separately in Caribbean and African ethnicities matched on age and household income. **A)** Distributions of age, ethnicity, and household income across Black ethnicities. **B)** AUC values were compared with unpaired DeLong tests. **C)** Mean absolute SHAP variable importances across Black ethnicities. Error bars indicate +/- one standard deviation (SD). A linear direction was calculated with a Pearson correlation on the averaged SHAP plot and is indicated by a light purple (feminine) or light green (masculine) color. Statistical differences between groups for each variable were calculated using ANOVAs, *p*-values were adjusted with a Bonferroni correction, and significant effects at $p<0.05$ are indicated with an asterisk.

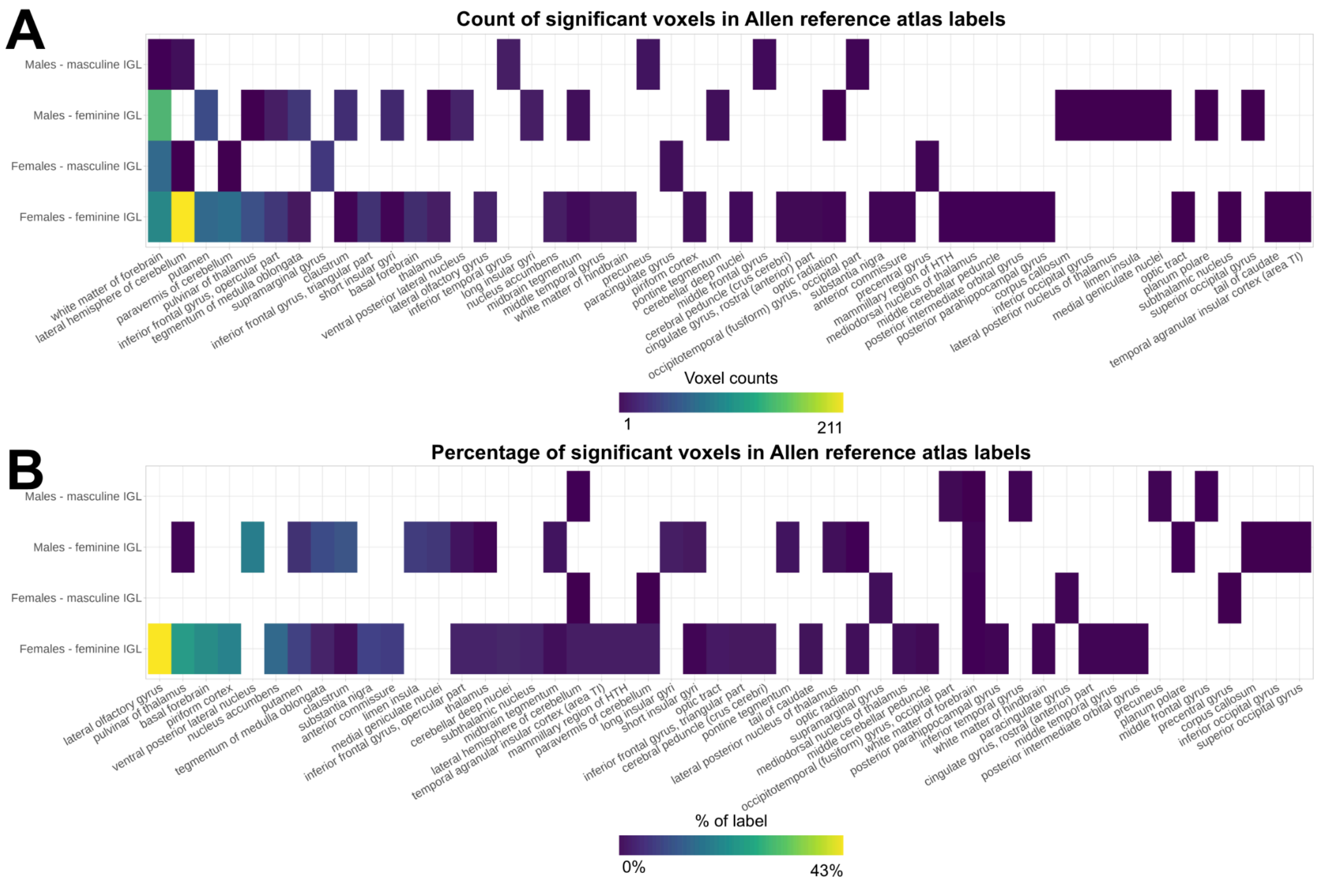

**Supplementary Figure 14. Overlap between significant voxels from deformation-based morphometry analysis and Allen reference atlas regions.** The number of significant voxels from the deformation-based morphometry analysis within each region of the Allen reference atlas was calculated separately for positive (larger with a masculine IGL) and negative (larger with a feminine IGL) effects in males and females. Results are presented **A)** with raw counts, and **B)** normalized for region size with the percentage of significant voxels relative to the total number of voxels of each region. Regions smaller than 10 voxels and regions without any significant voxels within them are not shown. Regions are sorted in descending order of the summed values across the four analyses.

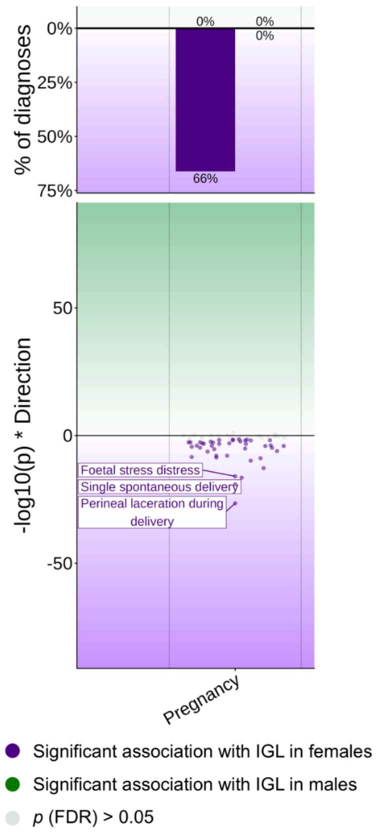

**Supplementary Figure 15. Associations between the IGL and pregnancy diagnoses, correcting for the number of live births.**

Pregnancy associations in Figure 5A were additionally corrected for the number of live births. We associated the IGL with medical diagnoses in males and females separately using logistic regression. Control groups were defined as individuals without any diagnoses in the pregnancy-related ICD-10 chapter. Significant effects at the FDR 5% level are shown. Top: the percentage of diagnoses showing significant effects in males and females (separately in the masculine and feminine direction) was calculated relative to the total number of diagnoses in the pregnancy ICD-10 chapter. Bottom: Miami plot of significant effects in males (dark green) and females (dark purple). The y-axis represents -$\log_{10}$($p$-values), with the feminine IGL direction indicated with negative values, and the axis limits are kept constant relative to Figure 5A. Non-significant effects are shown in gray. Notable diagnosis effects are labeled.

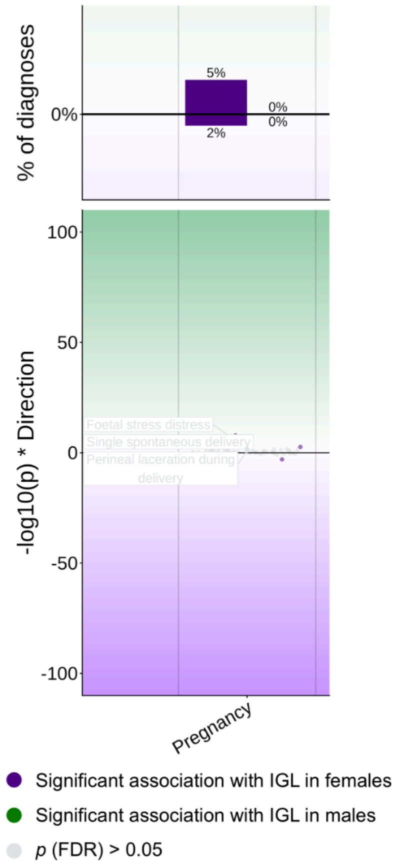

**Supplementary Figure 16. Associations between the Black IGL and pregnancy diagnoses, correcting for the number of live births.**

Pregnancy associations with the ethnicity-specific Black IGL were additionally corrected for the number of live births. We associated the IGL with medical diagnoses in males and females separately using logistic regression. Control groups were defined as individuals without any diagnoses in the pregnancy-related ICD-10 chapter. Significant effects at the FDR 5% level are shown. Top: the percentage of diagnoses showing significant effects in males and females (separately in the masculine and feminine direction) was calculated relative to the total number of diagnoses in the pregnancy ICD-10 chapter. Bottom: Miami plot of significant effects in males (dark green) and females (dark purple). The y-axis represents $-\log_{10}$(*p*-values), with the feminine IGL direction indicated with negative values, and the axis limits are kept constant relative to Extended Figure 10. Non-significant effects are shown in gray. Notable diagnosis effects are labeled.
